# Data-driven estimate of SARS-CoV-2 herd immunity threshold in populations with individual contact pattern variations

**DOI:** 10.1101/2021.03.19.21253974

**Authors:** Dan Lu, Alberto Aleta, Marco Ajelli, Romualdo Pastor-Satorras, Alessandro Vespignani, Yamir Moreno

## Abstract

The development of efficacious vaccines has made it possible to envision mass vaccination programs aimed at suppressing SARS-CoV-2 transmission around the world. Here we use a data-driven age-structured multilayer representation of the population of 34 countries to estimate the disease induced immunity threshold, accounting for the contact variability across individuals. We show that the herd immunization threshold of random (un-prioritized) mass vaccination programs is generally larger than the disease induced immunity threshold. We use the model to test two additional vaccine prioritization strategies, transmission-focused and age-based, in which individuals are inoculated either according to their behavior (number of contacts) or infection fatality risk, respectively. Our results show that in the case of a sterilizing vaccine the behavioral strategy achieves herd-immunity at a coverage comparable to the disease-induced immunity threshold, but it appears to have inferior performance in averting deaths than the risk vaccination strategy. The presented results have potential use in defining the effects that the heterogeneity of social mixing and contact patterns has on herd immunity levels and the deployment of vaccine prioritization strategies.

## 1 Introduction

The COVID-19 pandemic has forced the implementation of non-pharmaceutical interventions (NPIs) of different intensity with the aim of reducing the burden of the disease on the healthcare system and minimize deaths among the population. Preventive measures and severe restrictions alike strive for a reduction of social mixing and contacts among individuals ^1–7^, which help diminishing the transmission of the virus and ensure the proper working of health-care systems. Thus far, only a few countries have used NPIs to pursue a suppression policy with long and strict lock-downs until community transmission is locally eliminated followed by control at borders ^1^. The development of several efficacious vaccines against SARS-CoV-2 ^8, 9^, and the start of vaccination campaigns, open a new chapter in the fight against COVID-19. At the center of the discussion about vaccine is the so-called herd immunity threshold, i.e., when the epidemic starts to decline after a critical fraction of individuals in the host population is immune ^10, 11^. Herd immunity can be achieved either because infected hosts become immune once recovered or via vaccination of susceptible hosts. These two alternatives for the suppression of SARS-CoV-2 have been at the center of policy and public debates ^11^, along with the discussion concerning the calculation of the herd immunity threshold in realistic populations and transmission settings ^10, 12, 13^. Furthermore, the vaccine production constraints, the lack of definitive information on the sterilizing properties of the vaccines, and logistic issues in rolling out vaccination campaigns have raised the issue of what vaccination strategies should be implemented in order to avoid social disruption and avert the largest possible number of deaths regardless of herd immunity threshold considerations.

Here, we present a modeling framework to estimate herd immunity and vaccination coverage in populations with realistic individual contact pattern variations. We develop a multilayer representation of the social interactions of populations ^14^ from real data ^15^ gathered for several countries from all continents and use a SARS-CoV-2 stochastic transmission model ^16^ to estimate both the herd immunity threshold (HIT) and the overshoot infection level (OIL) resulting from the natural circulation of the virus. This approach allows the evaluation of different immunization strategies.

We find that vaccination policies that target first the more vulnerable groups (e.g., the elderly) require a higher vaccination coverage to reach herd protection than the ones designed to immunize individuals with more social mixing in the population -young and middle age adults-, but they have the potential to avert a significantly larger number of COVID-19 deaths even in the case of sterilizing vaccines. Our results allow framing the discussion of different strategies not only in the context of averted deaths, but also with respect to the overall prevalence of the disease in the population and the way forward to achieve suppression of SARS-CoV-2 in the population.

## 2 Methods

Assuming homogeneous mixing in the population, the HIT can be estimated using classical results of mathematical epidemiology and it depends only on the basic reproduction number (*R*_0_). Essentially, when the population is fully susceptible, a typical infectious individual generates on average *R*_0_ new infections. When the disease unfolds and more people get infected, the number of available susceptible persons decreases. Under these conditions, on average, only *R*_*t*_ = *R*_0_ · *s*_*t*_ of new infected persons per infected individual are produced, where *s*_*t*_ is the proportion of the population that is susceptible at time *t*. Thus, when *s*_*t*_ ≤ 1/*R*_0_, the effective reproductive number *R*_*t*_ goes below the epidemic threshold and an ongoing epidemic would start declining (inflection point). This leads to the well-known expression for the disease induced herd immunity threshold (DHIT) for homogeneously mixed populations DHIT= 1 − 1/*R*_0_ (as the proportion immune should be 1 − *s*_*t*_). Another quantity of interest is the disease-induced overshoot infection level (DOIL) that we identify with the level of immunity that is left in a population when an unmitigated epidemic is left unchecked ^17, 18^. The DOIL is generally considerably larger than the DHIT as, although the epidemic will start declining when the proportion of infected population is above the DHIT, the epidemic still generates new infections before dying out.

In a real population, however, there are multiple sources of heterogeneity that break down the homogeneous assumption when modeling the transmission of a pathogen like SARS-CoV-2 ^6, 15^. In this work, we focus on two important determinants for the evolution of the incidence of COVID-19 that are encoded in the structure of the population, namely, age and contact patterns. These two ingredients shape individuals’ social mixing, hence, how chains of transmissions emerge and grow. To this end, we build a multilayer network ^14, 19^, in which layers correspond to different age groups and connectivity patterns are encoded in both intra- and inter-layer connections. First, we collected age and behavioral mixing patterns for several countries that are structurally different in at least one of the two ingredients of the network model. Throughout the paper we will present the results for Italy, but we show in the SM the full results for several other countries from all continents. More specifically, we generate synthetic populations (*N* = 10^5^ individuals) and associate each individual to a node of the network. We assign to each node an age group and a number of contacts per unit of time, both extracted from empirical distributions characterizing the demographic structure and the behavioral (contact) pattern of the population (see the SM). We have considered 18 age-groups, which lead to a multilayer network made up by that same amount of layers, with a number of nodes per layer, *N*_*α*_, equal to the total number of persons of the age-group of layer *α* in the synthetic population.

To connect the nodes of the network while satisfying the age mixing patterns, several constraints must be verified. First, note that a property of the age-contact matrix *M* is its reciprocity, namely *M*_*αβ*_*N*_*α*_ = *M*_*βα*_*N*_*β*_, where *M*_*αβ*_ is the average number of contacts from an individual in age group *α* to individuals in age group *β*. Then, the probability that a node in the age group *α* is connected with any node in the age group *β* (including *β* = *α*) is *p*_*αβ*_ = *M*_*αβ*_/∑_*β*_ *M*_*αβ*_. Similarly, the probability that a node *i* chooses a node *j* of degree *k*_*j*_ situated in layer *β* to connect with is *k*_*j*_*/*∑_*l*∈*β*_ *k*_*l*_, where the denominator indicates the addition over the degree of all nodes present in layer *β*. Therefore, the likelihood that nodes *i* and *j*, of layer *α* and *β* respectively, are connected at each time-step is given by 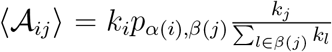, which satisfies the reciprocity property if and only if ∑_*l*∈*α*_ *k*_*l*_ = ∑_*β*_ *M*_*αβ*_*N*_*α*_, leading to the condition for the average degree on layer *α*, ⟨*k*⟩_*α*_ = ∑_*β*_ *M*_*αβ*_. Note that at each time-step we extract a new network realization from the ensemble generated by ⟨𝒜_*ij*_⟩. Fig. 1A schematically represents the resulting multilayer network for a single time-step ^19^.

**Figure 1:**
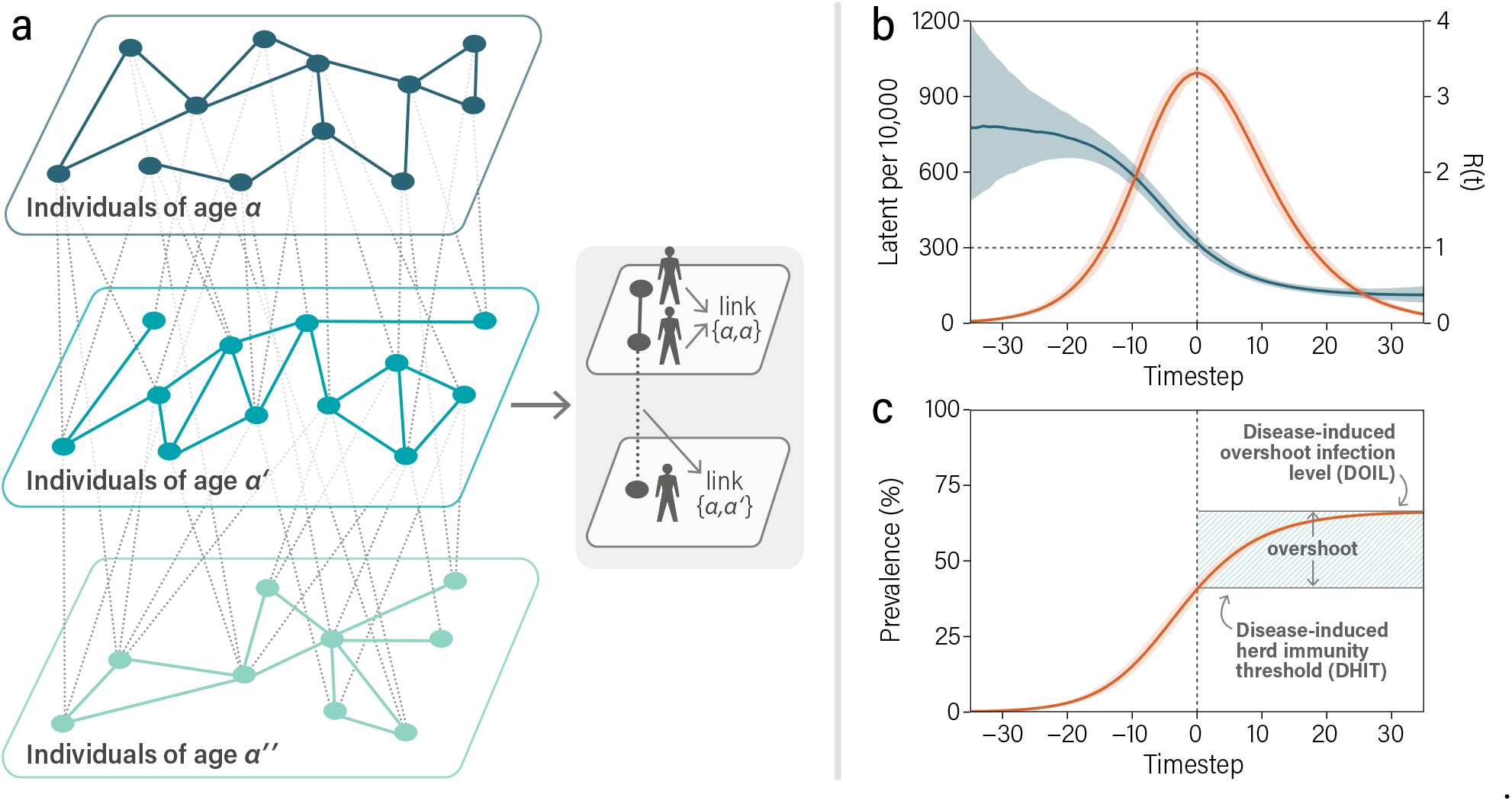
Model of population structure and herd immunity threshold. The structure of the population is encoded in a multilayer network, panel a, and consists of 18 layers, in which connections among nodes on the same layer correspond to mixing between individuals belonging to the same age group (diagonal elements of the contact matrix), whereas interlayer connections account for mixing between different age groups. Definition of DHIT, panels b and c: b) In orange, new latent individuals as a function of time. Simulations are temporally aligned so that the maximum number of latent individuals is reached a time equal to 0. In dark blue evolution of the effective reproduction number computed using the number of newly infectious individuals. We considered a gamma-distributed generation time with shape 2.12 and rate 0.32 (see the SM). c) Cumulative fraction of the population that has contracted the disease. We identify the DHIT and the DOIL as well as the overshoot fraction of infections.

The dynamics of the infection is described by a SEIR-like model that has been modified to accommodate COVID-19 natural history and the key aspects of SARS-CoV-2 transmission, including asymptomatic infectious individuals and several age-dependencies ^20, 21^ of the model parameters. Specifically, we implemented a stochastic, discrete-time compartmental model on top of the multilayer network in which individuals transition from one state to the other according to the distributions of key time-to-event intervals. Consequently, susceptible individuals (S) can be infected through contacts with either infectious symptomatic (IS), infectious asymptomatic (IA) or pre-symptomatic (PS) persons, progressing to the latent state (L), where they are infected but not infectious yet. Individuals in the latent compartment divert into two possible paths depending on whether they are symptomatic or not. On its turn, the former can develop diverse degrees of illness severity, leading to recovery or death. Additionally, we also take into account that the transmissibility depends on the age so that children and adolescents (under 20 y.o) have a susceptibility to the disease of 0.56 in comparison to adults ^20^. We incorporate this property in the first 4 layers of the network (as they contain the subset of individuals under 20 y.o.). A detailed description of the model and the parameters used is reported in the SM and in^16,22^. Our model does not allow a simple analytical calculation of the *R*_0_ as in the homogeneous assumption. In order to match our simulations with a specific *R*_0_ as measured in a real world setting, we consider the relationship between the reproduction number, epidemic growth rate, and generation time ^23^. In our analysis, we explore *R*_0_ values in the range 1.5 to 3.5. A full discussion of the *R*_0_ analysis is provided in the SM.

## 3 Results and Discussions

First, we consider the situation in which the infection circulates unmitigated through the host population (with no mitigation policies in place) and calculate the disease-induced herd immunity threshold (DHIT). This is defined as the proportion of all infected individuals at the inflection point (the maximum) in the curve of the incidence of latent individuals. This is the point in time at which the effective reproductive number for the generation of new infections (new latent individuals) goes below the epidemic threshold of 1 and the epidemic starts decaying, see Fig. 1B. The disease-induced overshoot infection level (DOIL) can then be calculated letting the system evolve till the epidemic dies out naturally because there are no more new infectious individuals circulating in the population, Fig. 1C.

We executed stochastic simulations of the epidemic transmission model and compute numerically the DHIT and DOIL values associated to the multilayer networks of the considered countries. In Fig. 2 we report the simulations for the multilayer network built on the Italian data. Our estimate for the disease-induced immunity level for the population of Italy in the case of *R*_0_ = 2.5 is 40.5% [95% CI 37.6-43.4]. This value represents the minimal proportion of the population that needs to acquire immunity through infection to thwart the circulation of the virus without any NPIs in place. In Fig. 2e we report the estimated DHIT and DOIL values for different *R*_0_ values (see Table 3 and 4 in the SM for the numerical values for *R*_0_ = 2.5). It is worth remarking that the obtained values differ considerably from the values obtained for the same *R*_0_ in a homogeneous model, because of the heterogeneous connectivity patterns of individuals.

**Figure 2:**
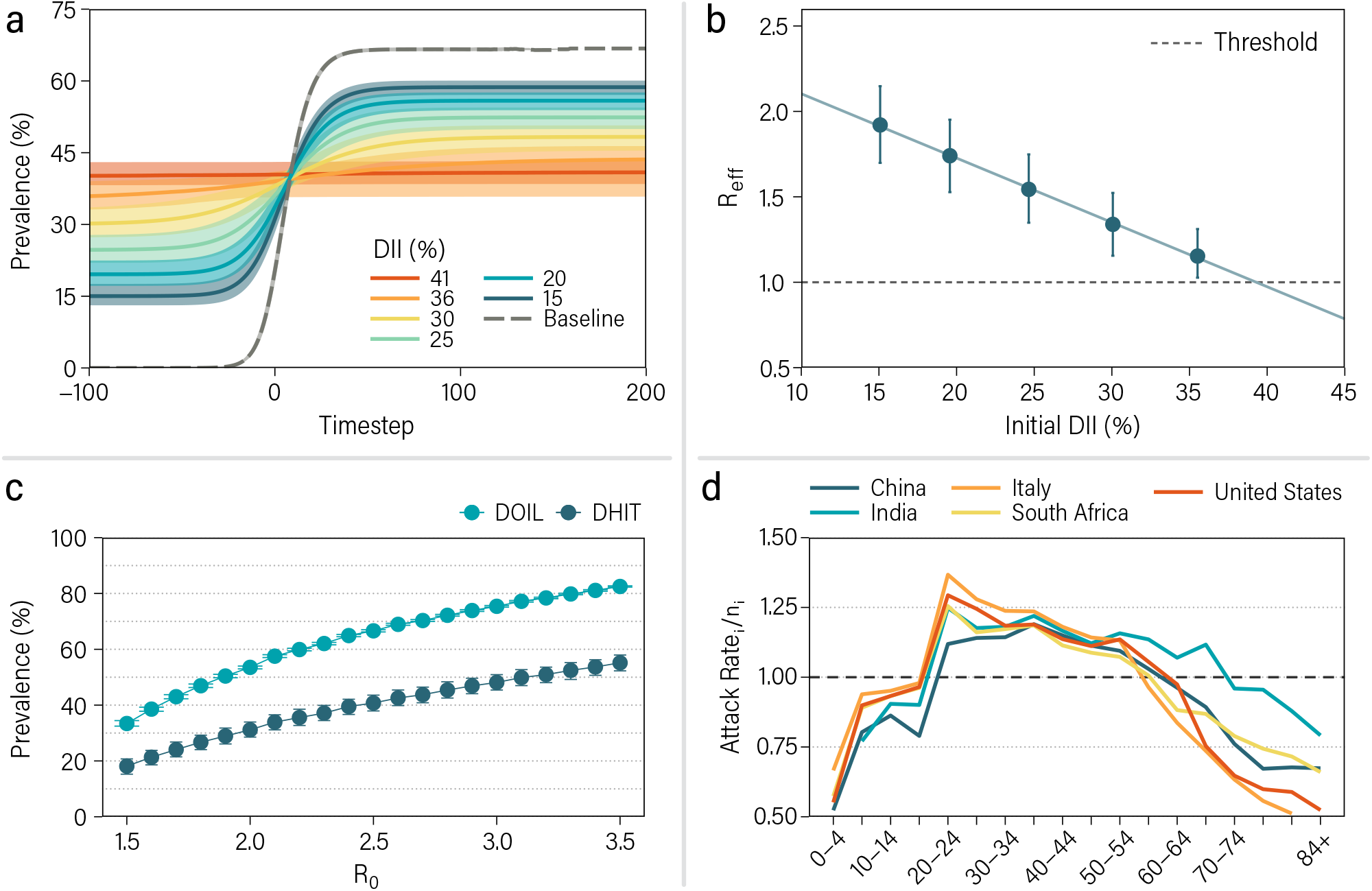
Disease-induced immunity threshold (DHIT). Disease-induced immunity and second waves, panels a and b: a) Prevalence (fraction of individuals who contracted the disease) in populations with different initial disease-induced immunity (DII). In all plots, solid lines represent the median value for 10.000 stochastic realizations and the shaded area the 95% C.I. b) The number of secondary infections produced by the seed individual, *R*_eff_, is measured in networks with different amounts of initial disease-induced immunity. Results shown in panels a and b correspond to the population of Italy and *R*_0_ = 2.5. Disease-induced immunity evolution, panels c and d: c) Prevalence in the Italian population for different values of *R*_0_ at the peak (DHIT) and at the end of the simulation (DOIL). d) Attack rate in each layer over the fraction of nodes that belong to that layer for different countries worldwide. Results in panel d) correspond are obtained for *R*_0_ = 2.5.

In order to simulate the epidemic behavior in a population that has achieved a specific disease-induced immunity level, we have performed further numerical simulations in which the system is evolved till a given proportion of the population is infected. Then, we use the disease-induced immune and susceptible population as the starting initial condition of a new epidemic with a single infected individual and record the DOIL reached by letting the epidemic spread unmitigated. This approach mimics a scenario in which the virus is circulating through the population until strict measures cut transmission down and reduce the incidence to vanishing values, which is followed by the reintroduction of new infectious persons once such restrictions are lifted. Fig. 2b shows the results obtained for several values of disease-induced immunity (DII) proportion when the pathogen is seeded again. As it can be seen, the DOIL depends on how close the system is to the DHIT. For the sake of comparison, we consider as the baseline the curve and DOIL corresponding to the situation in which the epidemic spreads unmitigated in a fully susceptible population. The closer the population is to the DHIT, the smaller the overshoot generated by a second epidemics and the final DOIL would be. It is important to stress that, upon reintroduction of infectious persons, immune individuals are not chosen at random, since their immune status is the result of having been infected during the course of the epidemics. If the same proportion of individuals were considered immune but selected at random, the previous history of the disease evolution would play no role and therefore the population would not be poised at DHIT. This is because the structure of the population induces dynamical correlations that determine which transmission paths are followed as the infection progresses. An important consequence of this result is that it highlights the impact that an intervention aimed at cutting transmission paths might have. Admittedly, if the community transmission is such that the incidence levels off to nearly zero locally, the acquired immunity of the population by natural infection could play a major role in suppressing local outbreaks once NPIs are withdrawn.

The previous observation can be further explored using a semi-analytical argument. We may define the number of secondary cases that the seed individual can produce upon reintroduction of the pathogen in the community as *R*_eff_ (in contrast to *R*_0_, which applies only for the first introduction to the community, i.e., when the whole population is susceptible). In classical heterogeneous populations this value is known to be dependent on both the mean and the variance of the number of contacts per individual ^24^. Actually, in the classical SIR model on networks, this value can be expressed as 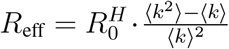, where 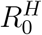 is the reproduction number in the homogeneous model and the second term is a function of the topological properties of the network, its average degree ⟨*k*⟩ and the second moment of the degree distribution, ⟨*k*^2^⟩^25,26^. In our model the previous history of the disease affects the terms depending on the moments of the degree distribution. Indeed, if some individuals have been removed from the susceptible population after a first wave of infections (either due to recovery or death), the structure of the network changes, modifying the value of *R*_eff_. If the network is in such state that *R*_eff_ < 1, then the reintroduction of the pathogen in the community cannot produce a large outbreak. However, if we remove nodes randomly the values of ⟨*k*⟩ and ⟨*k*^2^⟩ assume different values thus poising the population not at the DHIT. In Fig. 2c, we explore the value of *R*_eff_ as a function of the initial DII, while the results for the random selection of nodes are shown in Fig. 7 of the SM. Note that even though for a DII slightly smaller than 41% the average *R*_eff_ is close to the threshold, it is possible to have macroscopic outbreaks (see Fig. 6 of the SM).

The preceding expression highlights the role that the degree distribution has on the spreading of the disease. Because this distribution is partially determined by the age-mixing patterns of the population, it is to be expected that the path that the disease will follow should vary between regions. In section 5 of the SM we explore the values of DHIT and DOIL for different regions of the world. Since in our dataset the average degree in all regions is almost the same by construction, the differences in these values are small. Yet, in Fig. 2D we can see that the path followed by the disease is clearly different from region to region. This is revealed by looking at the attack rate within each age-group (layer) over the fraction of the population that belongs to that group at DHIT, which shows that some age-groups are more affected by the disease (values larger than 1) than what would be expected if the infections were randomly distributed across the population (or, in network terms, if there were no correlations between age and connectivity). This, in turn, has important implications towards the definition of vaccine prioritization strategies if we aim to reproduce the path the disease would follow in our population.

The above results are relevant to estimate what is the proportion of the population that needs to be vaccinated in order for the rest of susceptible individuals to be protected by the group immunity. Moreover, the vaccination coverage depends on how the population is vaccinated. We have considered three different scenarios for vaccination: *(i)* the classical random mass vaccination; *(ii)* a behavioral vaccine prioritization scheme in which the first individuals to be vaccinated are those that are more likely to transmit the disease; and *(iii)* a fatality-rate prioritization strategy that targets the eldest first and then vaccinates individuals in decreasing order according to their age. Specifically, at the initial state, a proportion *υ* of the population is immunized. We consider that the vaccine is 95% effective in preventing COVID-19 disease and forward transmission ^27^. In scheme *(i)*, the fraction *υ* of vaccinated individuals is chosen at random within the whole population. Scheme *(ii)* tried to mimic closely how disease-induced herd immunity is achieved. We first simulate the propagation of the disease up to the DHIT level. Then, we extract a random fraction *υ* of those individuals and vaccinate them. In this way, we mirror the way in which disease confers natural immunity targeting the individuals that would likely be infected during the course of the epidemic. Finally, in scenario *(iii)* the first to be vaccinated are those in the eldest group (85 y.o. or more) and the process continues down in age until the fraction *υ* of immune individuals is reached.

Fig. 3 shows the prevalence obtained when an initial proportion of the population is first vaccinated and then SARS-CoV-2 spreading is simulated in the population, for each of the strategies defined above. We have considered the same values of Fig. 2 for *υ* and a baseline scenario in which no individuals are immunized when the epidemic emerges. As expected, the prevalence of vaccinated populations decreases when the proportion of immunized individuals in the population increases. However, as not everyone in the population contributes equally toward the transmission of the infection, protecting the population as a whole is contingent on the vaccination strategy. Specifically, our results show that the best strategy in terms of reducing the prevalence is the second scenario, Fig. 3b, as our estimate for this scheme is consistent with the disease-induced threshold computed previously, i.e., a vaccination coverage close to the DHIT would protect the whole population from SARS-CoV-2 infection. Interestingly, random vaccination (Fig. 3a) also leads to lower prevalence values than the risk vaccination (Fig. 3c), as immunized individuals are spread through different age groups, which contributes to reducing the circulation of the virus with respect to the risk scenario, that only protects, for most values of *υ*, groups that in general play a secondary role in transmission. Indeed, in Fig. 3d we also show the final proportion of removed individuals (i.e., all individuals that are not susceptible at the end of the outbreak) for the different vaccination strategies and percentage of vaccinated population. Random and risk vaccination schemes lead to a final fraction of removed individuals that is greater than the DHIT, yielding comparable or greater levels with respect to the final prevalence in the baseline scenario. This also implies that the vaccine coverage needed for herd immunity with those strategies is generally considerably larger than the DHIT. The final proportion of infections instead depends on *υ* for the behavior-based vaccination that is mimicking the immunization introduced by the disease progression. Remarkably, although the risk strategy leads to the highest prevalence levels for any proportion of vaccinated individuals, it is the one that averts most deaths, even in the case of a vaccine that is extremely efficacious in blocking forward transmission. We estimate from simulations the total number of averted deaths for each of the vaccination strategies. Table 1 shows the number of averted deaths for each vaccine prioritization strategy with respect to the counterfactual unmitigated scenarios. Risk vaccination greatly reduces the number of deaths due to COVID-19 even for a proportion of vaccinated individuals as low as 15% -almost a factor 3 compared to the second best strategy. These results apply as well to vaccines with lower efficacy (see section 6 of the SM), although they require larger coverage for the behavioral strategy to work.

**Table 1:** Median number of averted deaths [95% C.I.] for each strategy per 10,000 individuals. In each strategy, the population is initially immunized through vaccination following the scheme explained in the main text. Results correspond to the population of Italy and *R*_0_ = 2.5.

| Number of averted deaths per 10,000 — $R_0 = 2.5$ | | | | | | |
| --- | --- | --- | --- | --- | --- | --- |
| Initially immune (%) | Random |  | Behavioral |  | Risk |  |
| 41 | 33.5 | [31.6 - 35.5] | 49.5 | [46.4 - 50.2] | 49.0 | [48.9 - 49.1] |
| 36 | 29.6 | [27.3 - 31.8] | 41.0 | [37.1 - 45.0] | 48.4 | [47.7 - 48.9] |
| 30 | 25.3 | [22.9 - 27.8] | 32.6 | [29.0 - 36.0] | 47.1 | [46.4 - 47.8] |
| 25 | 20.9 | [18.7 - 23.3] | 25.5 | [22.5 - 29.1] | 45.7 | [44.5 - 46.5] |
| 20 | 16.8 | [14.7 - 19.0] | 19.6 | [17.0 - 22.6] | 43.3 | [42.1 - 44.6] |
| 15 | 12.9 | [11.2 - 15.0] | 14.7 | [12.7 - 17.2] | 41.1 | [38.1 - 42.3] |

**Figure 3:**
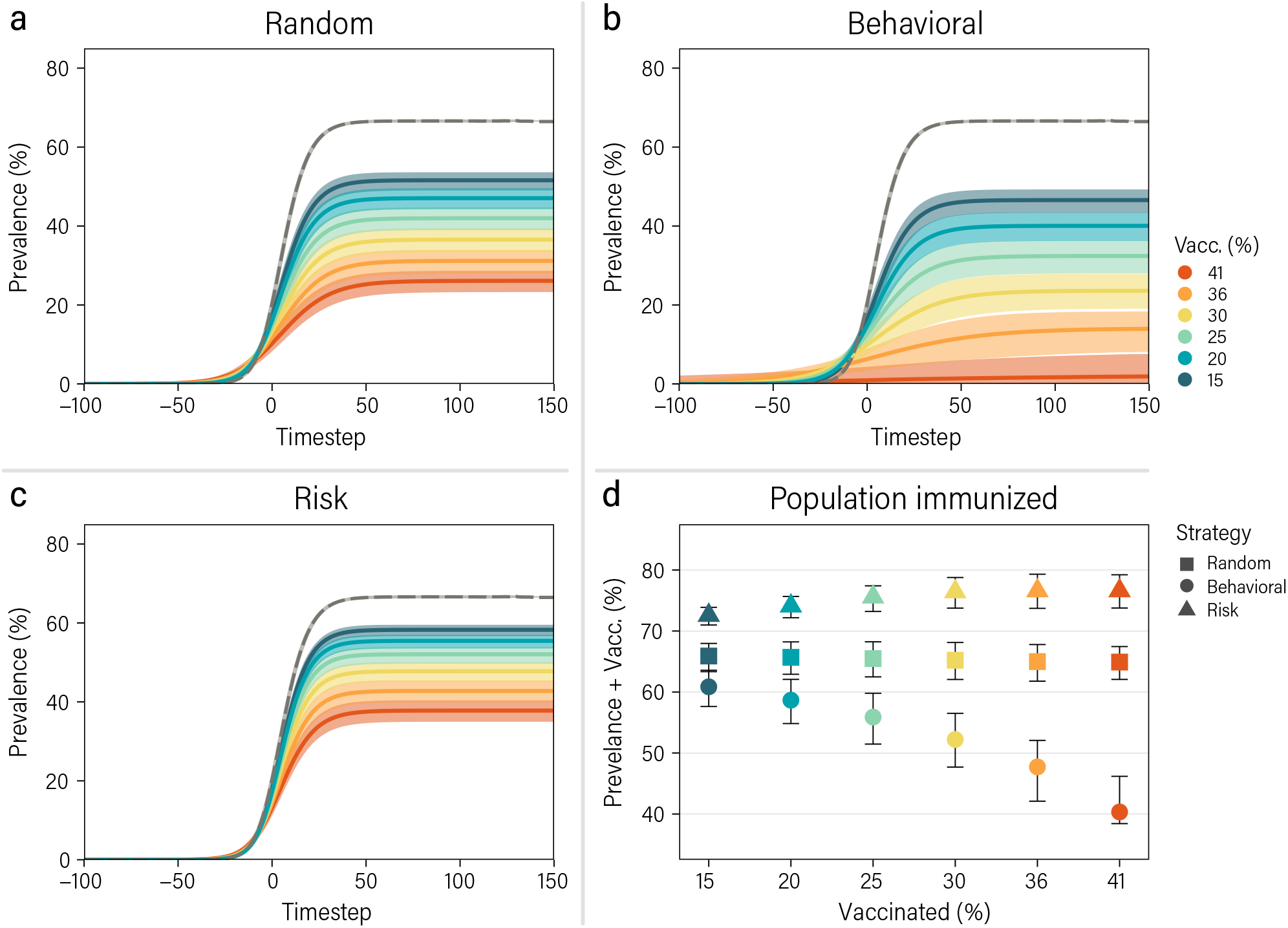
Effect of different vaccine prioritization strategies. Solid lines represent the median prevalence as a function of time under the random (a), behavioral-based (b) and risk-based (c) vaccine prioritization. The dashed lines account for a baseline scenario in which there is no vaccination nor any non pharmaceutical interventions. (d) Fraction of the population that is removed from the dynamics due to vaccine-induced immunity, disease-induced immunity or death once the disease dies out, for the different vaccination strategies as a function of the vaccine coverage. The results correspond to the population of Italy and *R*_0_ = 2.5.

These results were obtained under several assumptions. One important aspect is that we have considered idealized vaccine prioritization scenarios in which the temporal dimension has been neglected and also assumed a 100% adherence to vaccination. For instance, the former modeling choice hinders more realistic comparisons between the different strategies regarding how long the pandemic will be circulating in each scenario, the multiple dose administration, its social and economical implications ^28–32^. Likewise, adherence to vaccination ^33–35^ is important for higher values of coverage, in particular, to reach herd immunity levels of protection. We do not consider in our model the concurrency of the epidemic progression and the vaccination campaign and thus we do not address issues related to the timescale of the vaccination campaign roll-out. Differences in vaccine efficacy by age, vaccination schedule, and time from vaccine administration to partial and maximum protection were not considered as well as the epidemiological situation (e.g., natural immunity in the population at the start of the vaccination campaign, *R*_*t*_ during the vaccine roll out phase). Despite these simplifications, recent studies accounting for these aspects in age structured compartmental models recover results similar to those presented here ^36, 37^. Finally, it is worth stressing that we estimate the pre-pandemic DHIT, that is, we use contact matrices that capture the behavior of the population before any risk-avoiding reaction appears. However, it is known that individuals change their habits due to the presence of the disease and the enforcement of nonpharmaceutical interventions ^38–40^, and that behavioral changes do not stay constant along the full course of a pandemic. Therefore, the numerical estimates of the DHIT can vary as a result of behavioral changes.

## 4 Conclusions

Summarizing, in this work we have developed a data-driven multilayer population network that takes into account two factors that play a key role in shaping SARS-CoV-2 transmission and COVID-19 burden, namely, social mixing patterns as given by the number of contacts and age of the individuals of a population. Importantly, our framework only needs information at the population level, from which the network encoding the social mixing is built up. We provide numerical estimates of the DHIT and DOIL in realistic populations and show the effects of different vaccination strategies and rates on possible resurgences of the epidemic.

Our findings have the following important implications: (i) the variability in social interactions of the population determines the herd immunity threshold; it will thus be key to monitor the epidemiological situation as vaccines roll as people behavior and social contacts will change as well, potentially altering the herd immunity threshold and sparking new upsurge of COVID-19 cases; (ii) Risk-based and random vaccination scenarios require higher levels of vaccination coverage to suppress the circulation of SARS-CoV-2 as compared to the disease-induced herd immunity; it will thus be of paramount importance to extend vaccination effort well after reaching the theoretical herd immunity.

The previous implications suggest that more research is needed in order to find which is the optimal vaccination strategy considering, and possibly adapting to, the constantly evolving epidemiological situation and tailored to different populations as there is no one-fits-all solution. For instance, allowing for higher prevalence would keep health care systems under pressure for longer times, which in turn entails the need to keep restrictions and interventions. Additionally, it might have potential important consequences for virus evolution and the emergence of new variants as well as for public health systems given that the long term health consequences of suffering SARS-CoV-2 infection could be severe ^41^. Finally, our work also estimate that in all scenarios, except behavioral-based vaccine prioritization, the level of coverage needed to achieve herd immunity in the population is above some of the estimates of the proportion of the population that is willing to get vaccinated ^33, 35^, highlighting the need to nudge for higher vaccine acceptance.

## Data Availability

All data used is publicly available.

## Acknowledgements

Y.M. thanks M. Clarin for help with the design of Fig. 1. Y.M. acknowledges partial support from the Government of Aragon and FEDER funds, Spain through grant E36-20R (FENOL), and by MINECO and FEDER funds (FIS2017-87519-P). A.A. and Y.M. acknowledge support from Banco Santander (Santander-UZ 2020/0274) and Intesa Sanpaolo Innovation Center. M.A. and A.V acknowledge the support of the Cooperative Agreement number NU38OT000297 from The Centers for Disease Control and Prevention (CDC) and CSTE. RPS acknowledge financial support from the Spanish MICINN, under Project No. PID2019-106290GB-C21. D.L. acknowledges support from the China Scholarship Council though a PhD fellowship. The funders had no role in study design, data collection, and analysis, decision to publish, or preparation of the manuscript.

## Authors’ contributions

A.A., A.V., and Y.M. designed research; D. L., and A.A. performed research; D. L., A.A., M. A., R. P-S., A.V., and Y.M. analyzed the results. Y. M. wrote the first draft of the manuscript; D. L., A. A., M. A., R. P-S., A.V., and Y.M. discussed results and edited the manuscript. All authors approved the final version.

