## Supplementary material for "Data-driven estimate of SARS-CoV-2 herd immunity threshold in populations with individual contact pattern variations": SM file

**6 Vaccine efficacy. 18**

**7 Further vaccination scenarios. 23**

### 1 Network reconstruction

To build the multilayer contact networks we use the algorithm proposed in [1]. Given a node $i$ located in layer $\alpha$ and a node $j$ located in layer $\beta$, the expected number of links between nodes $i$ and $j$ is given by

$$\langle \mathcal{A}_{ij} \rangle = k_i p_{\alpha(i),\beta(j)} \frac{k_j}{\sum_{l \in \beta(j)} k_l} \tag{1}$$

where $k_i$ is the degree (number of links) of node $i$ and $p_{\alpha(i)\beta(j)}$ is the probability of establishing a link between a node in layer (age-group) $\alpha$ and layer $\beta$. This expression corresponds to the annealed version of the network. As such, in the simulation we randomly extract one realization of the network at each time-step so that the average of all of them corresponds to equation (1).

The value of $p_{\alpha,\beta}$ can be extracted from the mixing patterns matrix of the population as

$$p_{\alpha\beta} = \frac{M_{\alpha\beta}}{\sum_{\beta} M_{\alpha\beta}} \tag{2}$$

While the value of $k_i$, unless otherwise obtained, can be extracted from any degree distribution that we desire. Note, however, that the mixing matrix fulfills reciprocity, i.e.,

$$M_{\alpha\beta} N_{\alpha} = M_{\beta\alpha} N_{\beta} \tag{3}$$

where $N_\alpha$ is the number of nodes in layer $\alpha$. As such, equation (1) together with this last expression implies that

$$\langle k \rangle_{\alpha} = \sum_{\beta} M_{\alpha\beta}. \tag{4}$$

In other words, even though the degree distribution can be chosen freely, its average is fixed by the mixing matrix.

**1.1 Data.** The age mixing matrix of each region and country was obtained from [2]. The 85 age-groups originally considered in Ref. [2] were aggregated into 18 groups, going from age 0 to 84 in groups of 5 and a last group for 84 years old or older. For the degree distribution, we rely on the survey on contact patterns that was carried out in Italy for the POLYMOD project [3]. In such survey, the distribution of contacts per age-group can be described using a negative binomial distribution, see Fig. 1. For this reason, we choose a negative binomial distribution for the number of contacts per individual in each layer. As previously mentioned, the average of the distribution is fixed in each layer by the value of the age mixing matrix. However, the size of the distribution (also known as dispersion parameter) is not, since this parameter depends on the individual variability, but the matrices were obtained using data aggregated by age-group. To properly obtain the distribution, it would be necessary to carry out surveys similar to POLYMOD for each region under consideration, but this type of data is still scarce [4].

In this work, we have parameterized the size of the distributions based on the survey from Italy. This lack of data introduces a limitation for the comparison of different countries, since the different socio-cultural elements of each region might influence the variability of the distribution in some regions. In section 3.1, we explore two limits of the negative binomial distribution setting the dispersion parameter equal to (i) 1, obtaining a geometric distribution, and (ii) infinity, which yields a Poisson distribution.

### 2 SARS-CoV-2 transmission model

We use a stochastic, discrete-time compartmental model coupled to the multilayer contact network. The general structure of the model is depicted in Fig. 2. A susceptible individual ($S$) will become infected with probability $\beta_S$ if she contacts a pre-symptomatic ($P_S$) individual, $\beta$ if the contact is with an infectious symptomatic individual ($I_S$), and $r\beta$ if the contact is in the infectious asymptomatic state ($I_A$). Once infected, the individual will enter the incubation compartment ($L$) for a period extracted from a gamma distribution, $\epsilon$, during which she will be infected but not infectious yet. A latent individual will become infectious $\gamma$ days before the end of the incubation period, to account for pre-symptomatic transmission. Lastly, the individual will be removed (R) from the infectious pool according to an exponential process with rate $\mu^{-1}$, where $\mu$ is the average length of the infectious period in days. In order to check on the parameters' assumption we measured in the model the generation time that is in agreement with the epidemiological data. Note that the removed compartment does not imply recovery, only that the individual is no longer infectious. To estimate the number of deaths we later apply the empirical IFR to the set of removed nodes. When vaccination is taken into account a new compartment, $V$, is created, to distinguish between removed individuals who actually had the disease and those who did not.

In homogeneous populations, the basic reproduction number of this model can be expressed as:

$$R_0 = \frac{\beta r p}{(\gamma+\mu)^{-1}} + \frac{\beta(1-p)}{\mu^{-1}} + \frac{\beta_S(1-p)}{\gamma^{-1}} \tag{5}$$

where $\beta_S = \beta\gamma^{-1}k/\mu^{-1}(1-k)$. The description and values of all the parameters is shown in table 1. Note that this expression is only valid for homogeneous populations. In structured populations, the particular value of $R_0$ of each individual will depend on her connectivity. Furthermore, in layers 0 to 3, corresponding to age groups $[0-5), [5-10), [10-15), [15-20)$, individuals

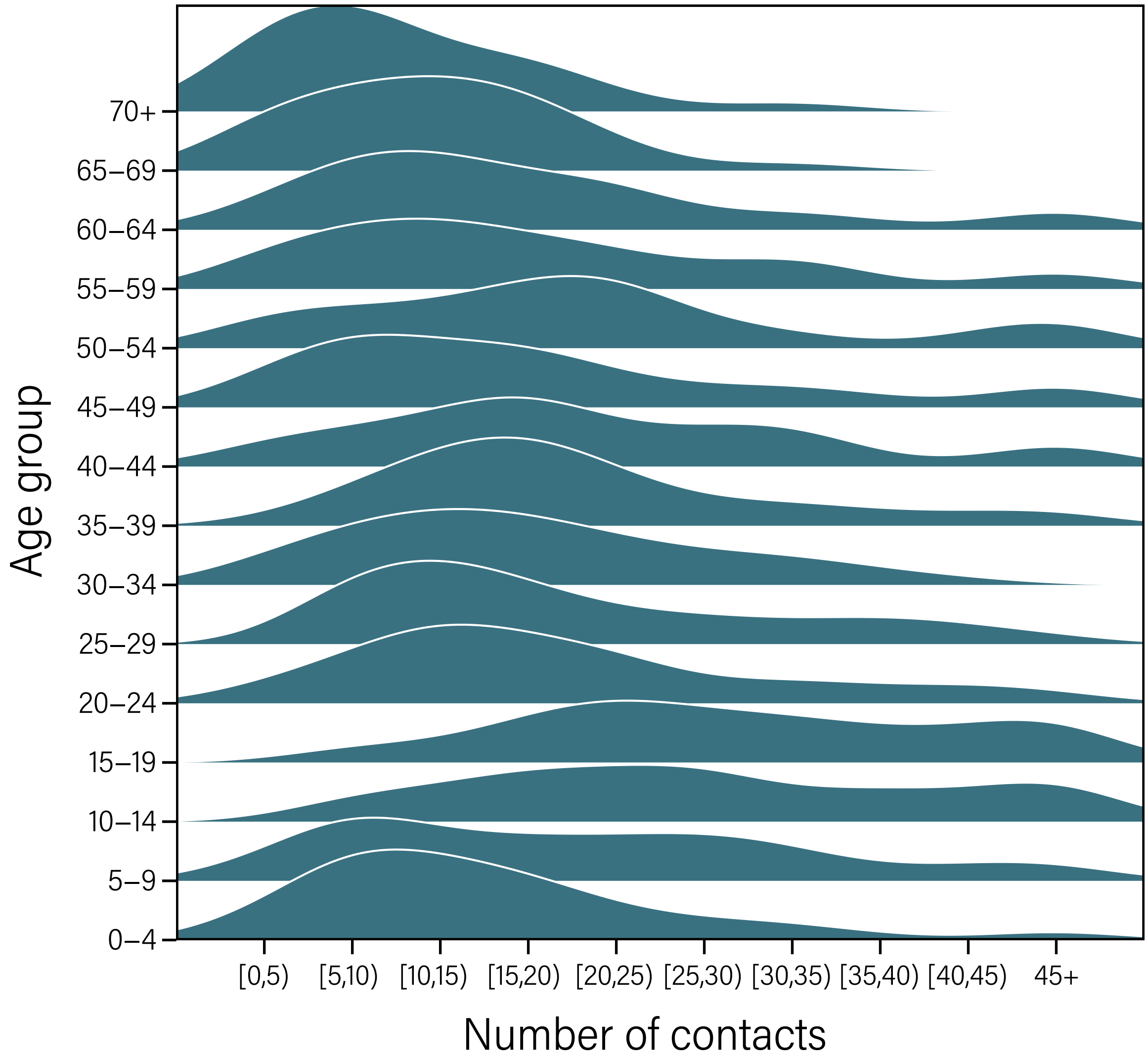

Figure 1: **POLYMOD survey**. Degree distributions for each age bracket measured using POLYMOD data for Italy. The maximum number of contacts that could be reported in the survey was 45, as such the empirical distributions are right-censored at that value.

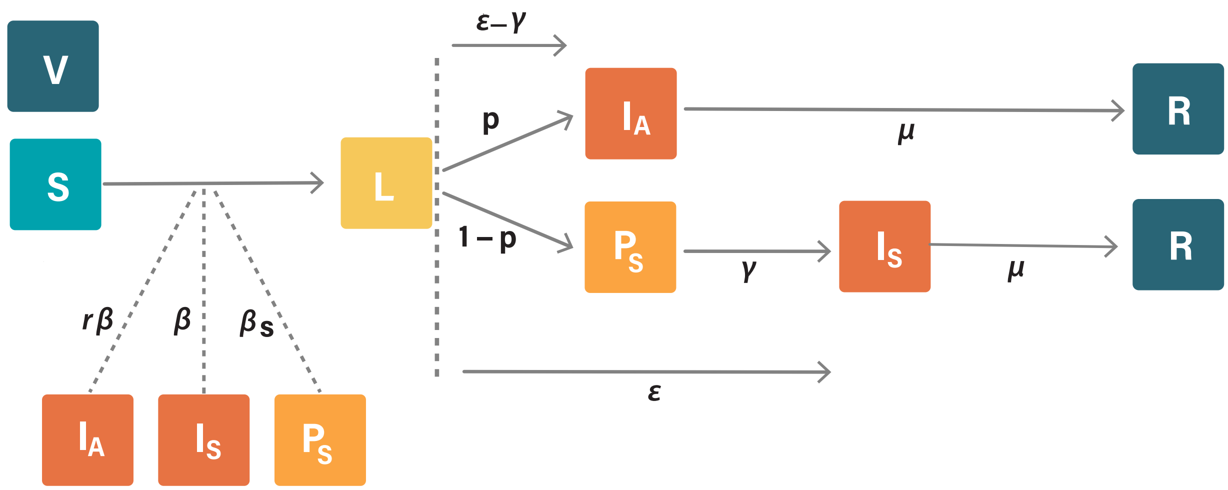

Figure 2: **Compartmental model description**. The different compartments and transition times from one to another are shown.

have a susceptibility to the disease of 0.56 [5]. As such, in each network, to select the appropriate value of $\beta$, we empirically estimate the dependency of $R_0$ with $\beta$. In figure 3 we show some of these distributions directly measured from the output of the model.

**2.1 Estimation of $R_0$.** In the classical SEIR model the value of $R_0$ can be estimated from the growth rate as $R_0 = (1 + rT_E)(1 + rT_I)$, where $i$ is the growth rate and $T_E$ and $T_I$ are the exposed and infectious periods, respectively. Since our model has a more complicated structure, we follow[10] and estimate the value of the basic reproduction number using the empirical (measured from the simulations) generation time. In particular,

$$R_0 = \frac{r}{\sum_{i=1}^{n} y_i (e^{-ra_{i-1}} - e^{-ra_i})/(a_i - a_{i-1})} \tag{6}$$

where $a_i$ are the category bounds in the histogram of the generation time and $y_i$ the observed relative frequencies. In Fig. 4, we show: (a) the temporal evolution of the prevalence from which the growth rate, $r$ can be estimated (b); (c) the generation time obtained from the simulation; and (d) the corresponding value of $R_0$.

| Parameters | Description | Age group | Value | Ref. |
|---|---|---|---|---|
| $r$ | relative infectiousness of asymptomatic individuals | - | 50% | † |
| $k$ | proportion of pre-symptomatic transmission | - | 50% | [6] |
| $\epsilon$ | incubation period (gamma distributed) | - | shape = 2.08<br>rate = 0.33 | [7] |
| $p$ | proportion of asymptomatic | - | 40% | [6] |
| $\gamma$ | pre-symptomatic period | - | 2 days | [8] |
| $\mu$ | time to removed | - | 2.5 days | $*$ |
| IFR | infection fatality ratio | 0-19<br>20-49<br>50-59<br>60-69<br>70-79<br>$\geq 80$ | 0%<br>0%<br>0.35%<br>0.88%<br>5.59%<br>8.15% | [9] |

Table 1: Set of parameters of the transmission model. †: assumed ;$*$: calibrated to the generation time $T_g$.

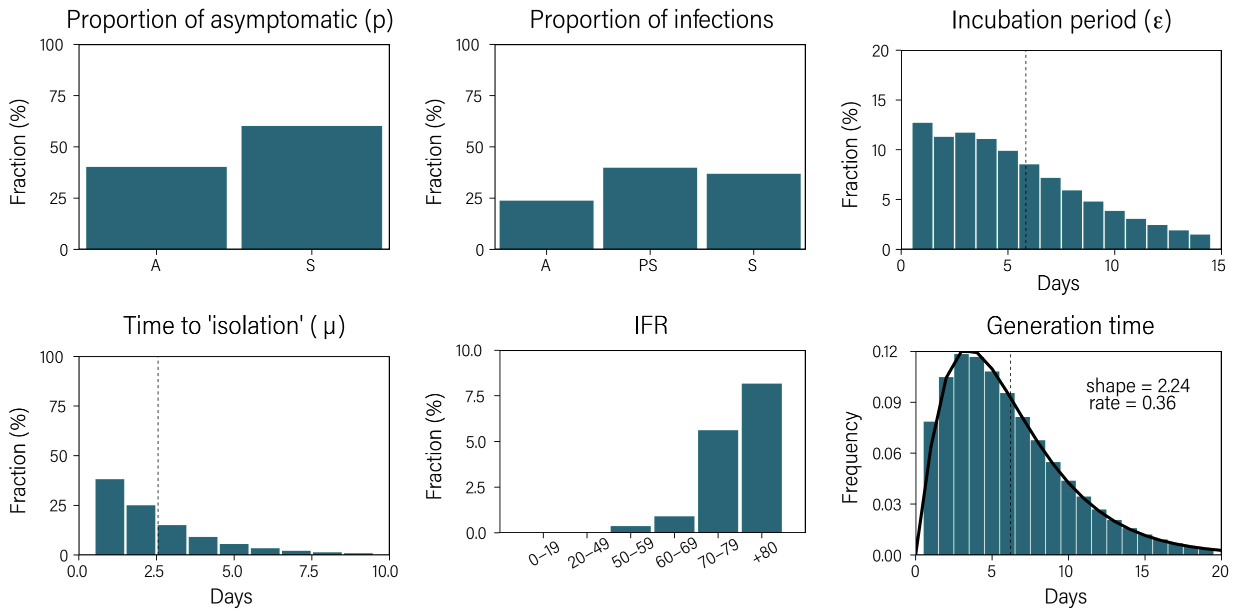

Figure 3: **Distributions obtained in the simulation.** Numerical distributions of the model parameters extracted from the simulations performed for Italy with $R_0 = 2.5$. The generation time distribution is well fitted by a gamma distribution with shape = 2.24 and rate = 0.36.

### 3 Further results on DHIT .

In Table 2 we report the numerical estimations of DHIT and DOIL for the Italian network.

**3.1 Influence of the degree distribution.** In a SIR-like model the scaling between the basic reproduction number in an heterogeneous network, $R_0^N$, and its homogeneous counterpart, $R_0^H$, can be expressed as:

$$R_0^N = R_0^H \frac{\langle k^2 \rangle - \langle k \rangle}{\langle k \rangle^2} \tag{7}$$

Hence, fixing $R_0^N = 2.5$ will yield a different value of $R_0^H$ depending on the heterogeneity of the network.

If we set the dispersion of the negative binomial distribution equal to 1, we recover the

| R0 | DHIT (%) | DOIL (%) |
|---|---|---|
| 1.5 | 18.1 [15.3-20.7] | 33.4 [32.5-34.5] |
| 1.6 | 21.3 [18.6-23.8] | 38.6 [37.8-39.3] |
| 1.7 | 24.1 [21.5-26.8] | 43.0 [42.3-43.7] |
| 1.8 | 26.7 [24.1-29.2] | 46.9 [46.3-47.6] |
| 1.9 | 28.9 [26.1-31.8] | 50.4 [49.8-51.0] |
| 2.0 | 31.1 [28.5-33.9] | 53.5 [53.0-54.0] |
| 2.1 | 33.9 [31.1-36.5] | 57.5 [57.1-58.1] |
| 2.2 | 35.6 [32.7-38.5] | 59.9 [59.4-60.4] |
| 2.3 | 37.1 [34.5-39.9] | 62.1 [61.6-62.5] |
| 2.4 | 39.5 [36.6-42.1] | 64.9 [64.5-65.3] |
| 2.5 | 40.5 [37.7-43.5] | 66.3 [65.9-66.7] |
| 2.6 | 42.6 [39.8-45.4] | 68.9 [68.6-69.3] |
| 2.7 | 43.7 [40.8-46.5] | 70.3 [70.0-70.7] |
| 2.8 | 45.4 [42.5-48.4] | 72.2 [71.9-72.5] |
| 2.9 | 46.9 [44.1-49.6] | 73.9 [73.6-74.2] |
| 3.0 | 48.1 [45.4-50.9] | 75.4 [75.1-75.7] |
| 3.1 | 49.9 [47.0-52.7] | 77.2 [76.9-77.5] |
| 3.2 | 50.9 [48.0-53.6] | 78.4 [78.1-78.7] |
| 3.3 | 52.4 [49.2-55.2] | 79.8 [79.6-80.1] |
| 3.4 | 53.7 [50.7-56.3] | 81.1 [80.8-81.4] |
| 3.5 | 55.2 [52.3-58.0] | 82.5 [82.3-82.7] |

Table 2: Estimated disease-induced herd immunity threshold (DHIT) and disease-induced overshoot infection level (DOIL) as a function of $R_0$ for the network of Italy. DHIT is measured as the median prevalence [95% C.I.] over 10,000 runs at the peak of latent individuals, while DOIL is measured as the median prevalence at the end of each run. Note that these values are estimated on the same annealed network, so that we implicitly assume that individuals do not change their behavior due to the disease. In an actual scenario, the introduction of non-pharmaceutical interventions and the change on the behavioral patterns of the population will modify the interaction network and, thus, change the local value of DHIT and DOIL. Nevertheless, if DHIT or DOIL is temporarily achieved under a modified interaction network, once the behavior of the population goes back to normal the values of DHIT and DOIL should get close to this baseline scenario.

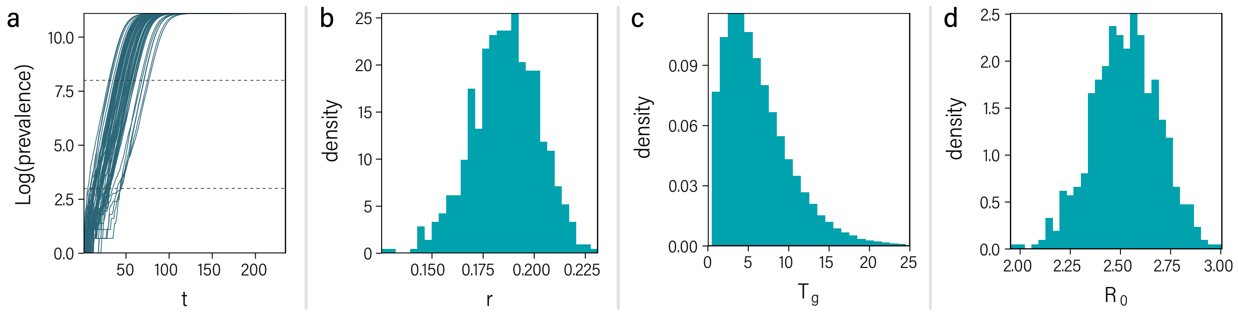

Figure 4: **Estimation of $R_0$.** a) Temporal evolution of the logarithm of the prevalence. b) Estimated growth rate. c) Generation time extracted from the simulation. d) $R_0$ obtained using equation (6).

gamma distribution. Expression (7) is thus

$$R_0^N = 2R_0^H \tag{8}$$

regardless of the average of the distribution. Similarly, if we take the upper limit of the dispersion (infinity) we obtain a Poisson distribution which yields

$$R_0^N = \left(1 + \frac{2}{\lambda}\right) R_0^H \tag{9}$$

where $\lambda$ is the mean value of the distribution.

If we fix $R_0^N = 2.5$ in all three cases we obtain $R_0^H(\text{Gamma}) = 1.25$, $R_0^H(\text{Negative Binomial}) = 1.71$ and $R_0^H(\text{Poisson}) = 2.14$. In an homogeneous population the final size of an epidemic can be expressed as

$$\ln \frac{s_0}{s_\infty} = R_0 \left[1 - s_\infty\right] \tag{10}$$

which can be solved numerically to obtain the theoretical prevalence at the end of the simulation as $1 - s_\infty$. In particular, we obtain a final prevalence of 40%, 70% and 83% for the gamma, negative binomial and Poisson distributions. In Fig. 5, we show the results for these three distributions.

These results highlight the importance of properly characterizing the way in which societies interact, since empirical estimations of $R_0$ based on surveillance data always include implicitly the effect of the network. Thus, if the heterogeneity of the interactions is unknown, the range of possible outcomes is too wide.

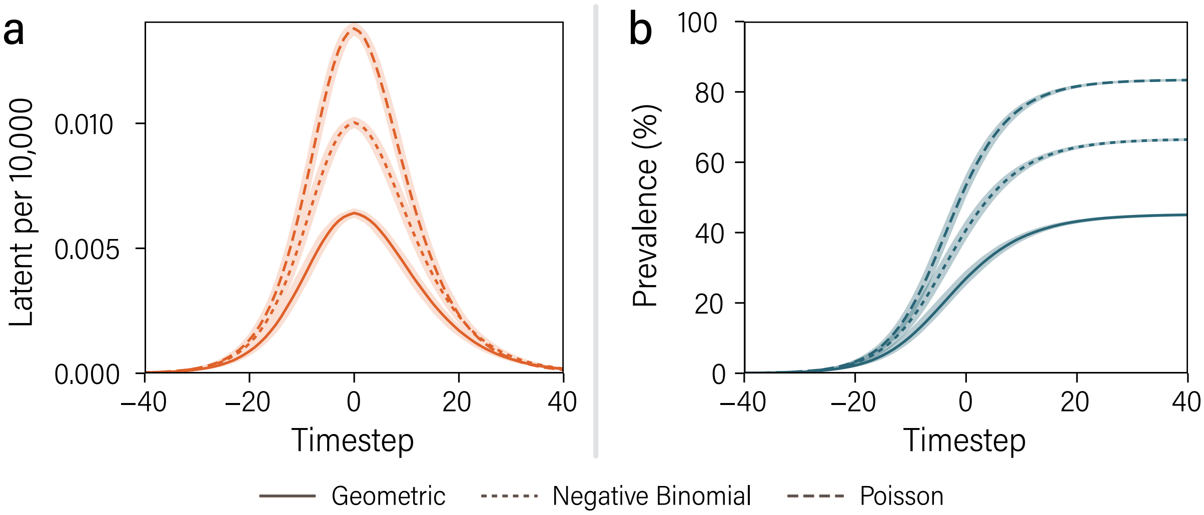

Figure 5: **Effect of degree distribution**. Median value of the amount of latent individuals (left) and prevalence (right) together with the 95% C.I. (shaded regions). In all cases the degree distribution follows a negative binomial distribution with dispersion: equal to 1 (yielding a geometric distribution, solid lines); equal to infinity (Poisson distribution, dashed lines); and as extracted from the POLYMOD data (dotted lines).

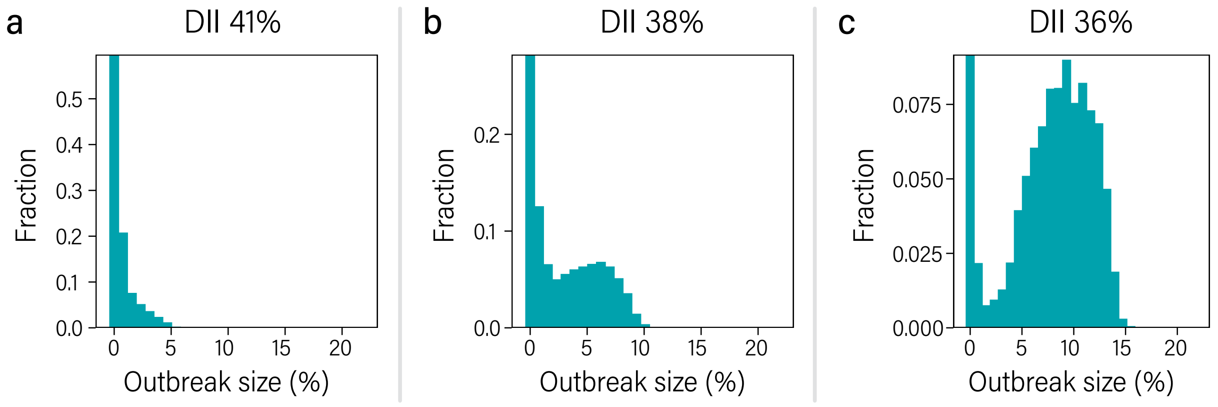

Figure 6: **Outbreak size in DII**. Size of outbreaks produced with different values of initial disease induced immunity in the population. Panel a shows the results for an initial immunity at the DHIT value, while b and c show examples of situations with slightly smaller initial immunity.

**3.2 Outbreaks after reaching DHIT through DII.** In Fig. 6, we study the size distribution of outbreaks produced in a hypothetical second introduction of the pathogen in populations that completely eliminated it within the DII scenario. We start each run with the whole population in the susceptible state. After a certain fraction of the population has been infected, we completely stop the spreading - simulating a very strict individual-based lockdown scenario - after all infected individuals move to the removed compartment. Afterwards, we introduce again the pathogen in a random seed. Since it is possible that the seed is weakly connected to the system, due to the removal of the nodes that were infected in the previous wave, sometimes it will not produce a second outbreak. Hence, we repeat this process 1,000 times for each run, and store only those outbreaks that spread, at least, to more than 100 nodes (0.1% of the system). In panel a, we show the fraction of events leading to an outbreak of a given size when the first wave infects 41% of the population (corresponding to the DHIT value). In panels b and c we do the same but with 38% and 36% of initial disease induced immunity, respectively. Clearly, while outbreaks in the situation close to DHIT are very small, once we move away from that point a second peak in the distribution appears, reflecting the emergence of a giant component in the network created by the disease path.

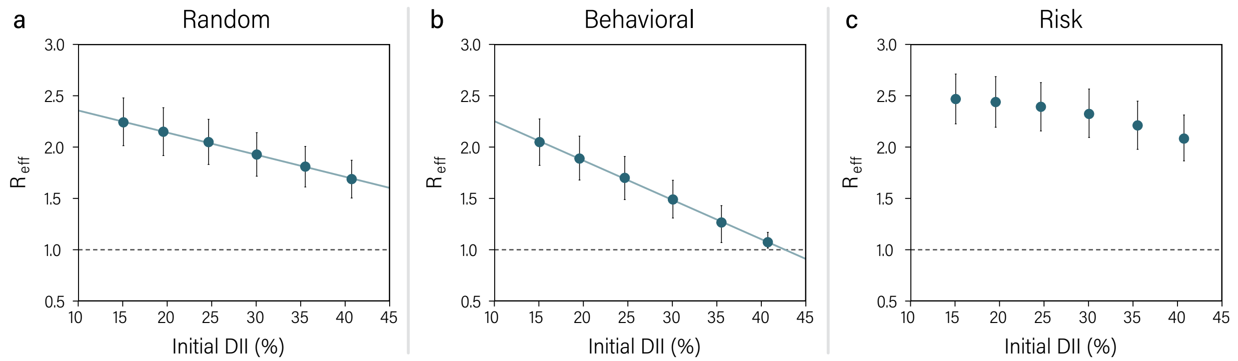

Figure 7: **Epidemic threshold and vaccination**. Effective reproductive number in the Italian network after the vaccination of a certain amount of individuals following the random (a), behavioral (b) or risk (c) vaccine prioritization strategies.

### 4 Epidemic threshold with vaccination.

The same approximation used in the main text to define the epidemic threshold in the DII scenario can be applied to the networks with vaccination. To do so, we first vaccinate the corresponding amount of individuals according to the chosen strategy and efficacy. Then, we measure $R_{\text{eff}}$ and compare it with the threshold. The results, Fig. 7, show how $R_{\text{eff}}$ is modified as a function of the vaccination coverage. In terms of network topology, in the random strategy it is possible that some of the largest connected nodes - also known as hubs - get vaccinated, lowering $\langle k^2 \rangle$ and, hence, $R_{\text{eff}}$. However, the behavioral strategy is much more effective in this regard, since vaccination is focused on a set of nodes that is more likely to have hubs. On the other hand, the risk strategy focuses on age groups that do not have any hubs, and hence it barely modifies the value of $R_{\text{eff}}$ until relative large values of the coverage, when it reaches age groups that do have hubs.

### 5 Impact of socio-cultural and environmental factors.

Socio-cultural factors can influence the transmission in our model in 3 ways: (i) by changing the degree distribution in each layer; (ii) by modifying the shape of the age mixing patterns; and (iii) through cultural or environmental factors that influence how effective is the transmission of the virus, i.e., the value of $\beta$ in our model.

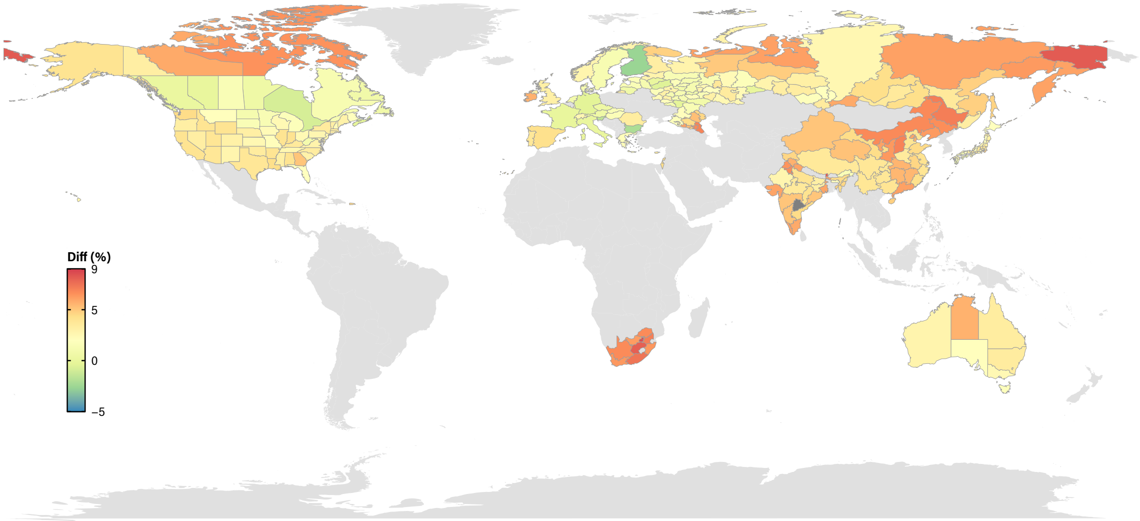

Figure 8: **DHIT in each region with fixed** $R_0$. Relative difference of the DHIT value obtained in each region in comparison with the one in Italy. We set $R_0 = 2.5$ and a dispersion parameter for the distributions equal to the one in Italy

.

Unfortunately, we cannot analyze the differences induced by the first factor since that would require precise information on the individual variability of people in each region, something that is not currently available as discussed in section 3.1. The second factor, on the other hand, can be easily studied thanks to the availability of high resolution mixing matrices [2]. In Fig. 8, we present the value of DHIT obtained in each available region with $R_0 = 2.5$ in comparison to the one obtained for Italy. The absolute DHIT values for each country are presented in table 3.

The previous factors, in essence, only modify the contact matrix. However, there are multiple other elements that can modify the transmissibility of the virus: the way in which individuals of a certain culture greet each other, the distance at which individuals usually interact, the way in which they talk, whether they are used to wear masks, as well as environmental factors that could enhance or diminish the spreading of the virus. In terms of our model, this would imply a different value of $\beta$ for each region which, once applied to each network (i.e., population structure), could in turn yield a larger or smaller $R_0$. Note that $\beta$ and $\langle k \rangle$ are not necessarily correlated, and there could be the case of regions with larger $\beta$ and lower $\langle k \rangle$ having the same value of $R_0$ as regions

| Country | DHIT (%) | | Country | DHIT (%) | |
|---|---|---|---|---|---|
| Australia | 41.4 | [38.8 - 44.0] | Italy | 40.5 | [37.7 - 43.5] |
| Austria | 40.6 | [37.5 - 43.5] | Japan | 41.6 | [38.6 - 44.4] |
| Bulgaria | 39.6 | [36.7 - 42.4] | Latvia | 41.0 | [38.0 - 43.7] |
| Canada | 40.8 | [37.5 - 43.8] | Lithuania | 40.7 | [37.5 - 43.4] |
| China | 42.7 | [39.7 - 45.8] | Luxembourg | 41.2 | [38.5 - 44.1] |
| Cyprus | 41.8 | [39.0 - 44.8] | Netherlands | 40.8 | [37.9 - 43.8] |
| Czech | 40.6 | [37.7 - 43.1] | Norway | 41.5 | [38.8 - 44.4] |
| Denmark | 40.9 | [37.9 - 43.6] | Portugal | 41.9 | [38.7 - 44.7] |
| Estonia | 41.1 | [38.0 - 44.0] | Romania | 41.8 | [38.6 - 44.7] |
| Finland | 39.4 | [36.4 - 42.3] | Slovakia | 41.0 | [38.0 - 43.7] |
| France | 40.4 | [37.4 - 43.0] | Slovenia | 39.8 | [37.1 - 42.6] |
| Germany | 40.3 | [37.2 - 43.0] | South Africa | 43.6 | [40.3 - 46.5] |
| Greece | 41.3 | [38.4 - 44.0] | Spain | 42.1 | [39.4 - 45.0] |
| Hungary | 41.1 | [37.9 - 43.8] | Sweden | 40.9 | [38.0 - 43.5] |
| India | 42.4 | [39.8 - 45.2] | Switzerland | 41.5 | [38.7 - 44.2] |
| Ireland | 42.7 | [39.7 - 45.3] | United States | 41.6 | [38.9 - 44.3] |
| Israel | 42.2 | [39.5 - 45.3] | United-Kingdom | 41.7 | [38.7 - 44.6] |

Table 3: DHIT value in each country with $R_0 = 2.5$.

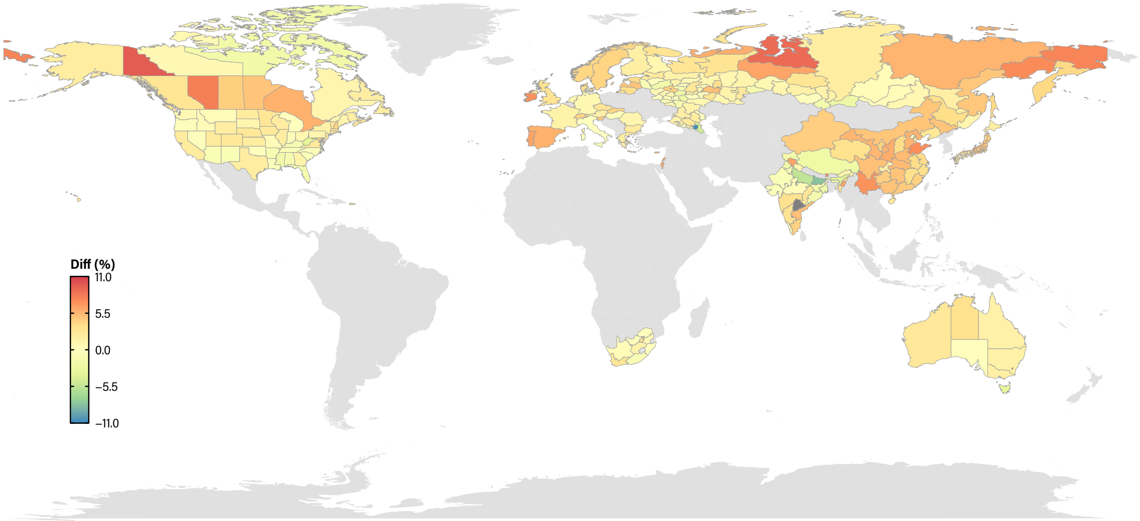

Figure 9: **DHIT in each region with fixed** $\beta$. Relative difference of the DHIT value obtained in each region in comparison with the one in Italy. We set the value of $\beta$ in all regions to the one that yields $R_0 = 2.5$ in Italy.

with lower $\beta$, as long as their $\langle k \rangle$ were larger. In fact, in the previous comparison we fixed $R_0$ in all regions, so that the value of $\beta$ was adapted to the specific contact matrix.

Conversely, in Fig. 9, we fix the value of $\beta$ to the one that sets the value of $R_0$ equal to 2.5 in Italy, and apply it to the rest of the regions. Hence, the value of $R_0$ will vary, being larger in those areas with larger $\langle k \rangle$, and smaller in those with a smaller $\langle k \rangle$. In this case, we observe larger differences, up to $8\%$. However, note that in the extraction of the mixing matrices it is assumed that the number of contacts in each setting is the same, regardless of the country (see [2] for details). Thus, if more precise information on the degree distributions in each country was available, the divergences could be much higher. In figures 10, 11 and in table 3 we report the corresponding values for DOIL as well.

**5.1 Age-groups attack rate and DHIT.** The fact that the DHIT and DOIL values are similar in all settings is a consequence of having almost the same average degree in all regions, which is rooted into the assumptions behind the construction of the mixing matrices. However, the shape of the matrices is determined by the socio-cultural characteristics of the population and, as such, it

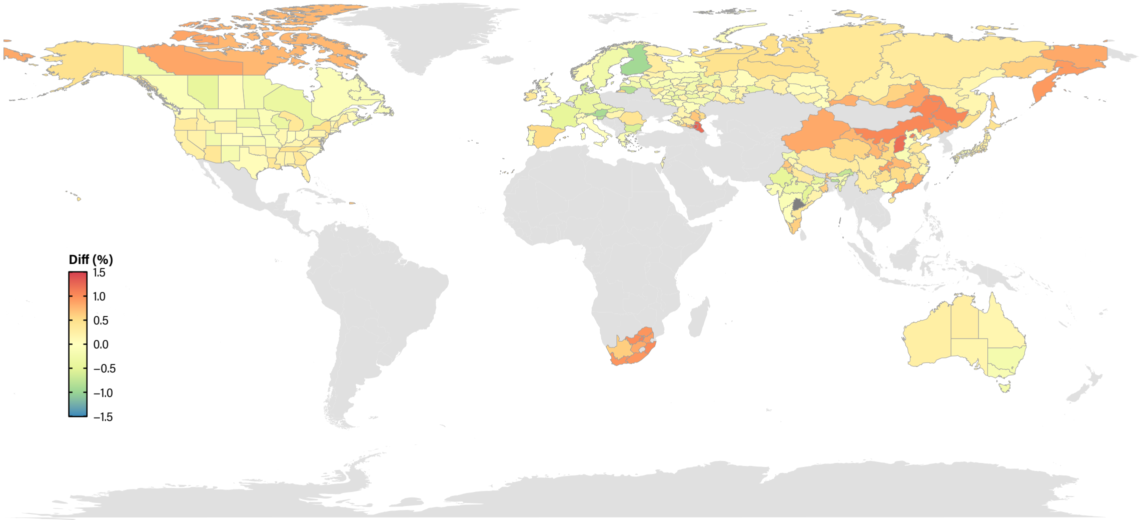

Figure 10: **DOIL in each region with fixed** $R_0$. Relative difference of the DOIL value obtained in each region in comparison with the one in Italy. We set $R_0 = 2.5$ and a dispersion parameter for the distributions equal to the one in Italy

.

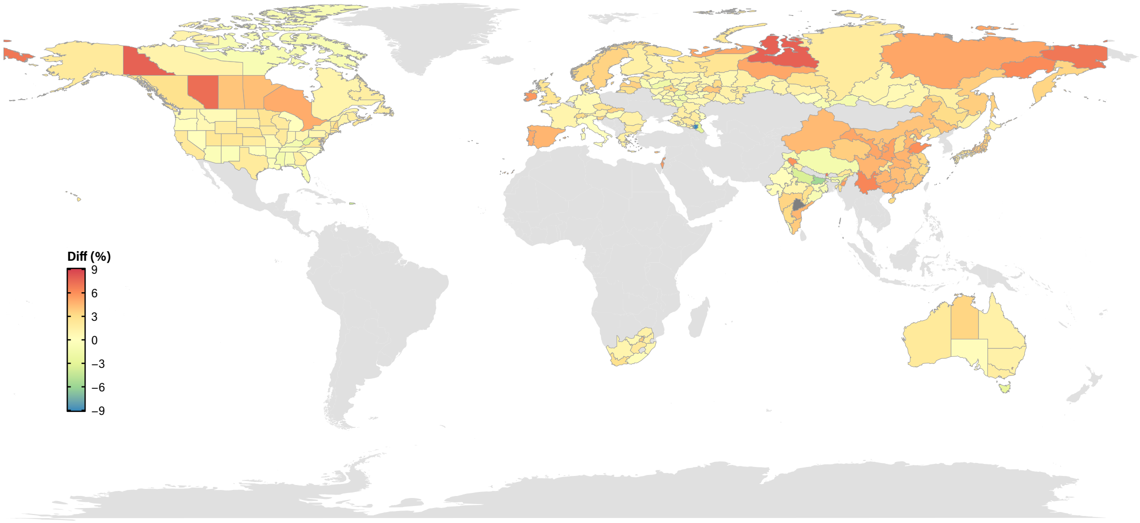

Figure 11: **DOIL in each region with fixed** $\beta$. Relative difference of the DOIL value obtained in each region in comparison with the one in Italy. We set the value of $\beta$ in all regions to the one that yields $R_0 = 2.5$ in Italy.

varies from region to region. Hence, even though the amount of individuals who got the infection at DHIT might be the same, their demographics will depend on the region.

Indeed, in Fig. 12 we show the attack rate within each layer over its relative size. If nodes were distributed randomly across layers or, in other terms, age and connectivity were completely uncorrelated, that quantity should be 1 for all age groups. Yet, we observe that certain age-groups are much less affected than it would be expected, with values lower than 1, and others much more. In particular, the disease tends to spread less within the layers containing children due to their lower susceptibility to the disease. On the other hand, young-adults are much more affected by the disease. Lastly, the elderly are less affected mainly because they are excluded from the workplaces and school settings. Since these two groups - young-adults and the elderly - are the ones further apart, in Fig. 13 we show the difference in their attack rate proportion, i.e.

$$\frac{\text{AR}_{\text{young-adults}}}{n_{\text{young-adults}}} - \frac{\text{AR}_{\text{elderly}}}{n_{\text{elderly}}}$$

### 6 Vaccine efficacy.

In the results shown in the main text, it was assumed that the vaccine has an efficacy of 95% in preventing forward transmission and severe illness. In this section, we explore the results when the efficacy is 85% and 60%, see Figs. 14-15 and Tables 7 and 8. In both cases, we observe that the best strategy is the one based on the infection fatality risk, which is to be expected given the dynamics of the system. The disease-induced immunity threshold was calculated to be at 41% for the Italian population. Immunizing a similar amount of nodes using a vaccine with high efficacy while targeting the individuals that are more likely to be in the path of the disease - the behavioral scenario - aims at mimicking such a situation. However, if the efficacy is not that high, it is clear that it will be necessary to vaccinate a larger amount of nodes in order to reach the required 41% of actual immune individuals. This is why the behavioral strategy with only 60% or 85% of vaccine efficacy is not able to thwart the spreading of the disease, since the DHIT has not been reached yet. On the other hand, the risk strategy directly shields those individuals that are more likely to die, at the cost of not controlling the propagation of the disease. This, in turn, grants a larger amount of averted deaths. Thus, the results show that for the behavioral strategy to work with a vaccine with

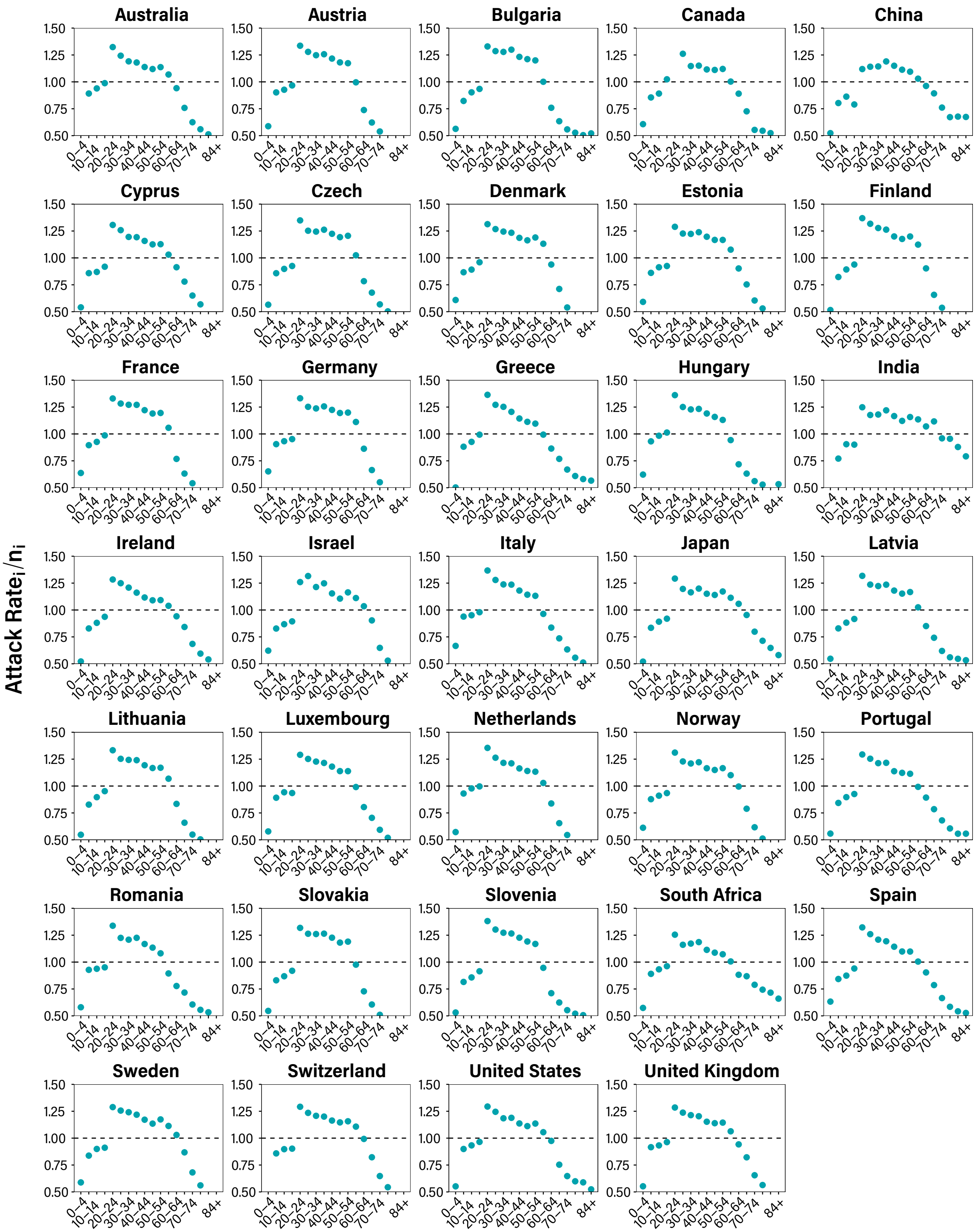

Figure 12: **Age distribution of infected individuals at DHIT**. The $y$-axis shows the attack rate of each age group (layer) over the relative size of the layer. In all cases, $R_0 = 2.5$.

| **Country** | **DOIL (%)** | | **Country** | **DOIL (%)** | |
|---|---|---|---|---|---|
| Australia | 67.4 | [67.0 - 67.8] | Italy | 66.3 | [65.9 - 66.7] |
| Austria | 66.4 | [65.9 - 66.7] | Japan | 67.8 | [67.4 - 68.2] |
| Bulgaria | 65.0 | [64.7 - 65.5] | Latvia | 67.1 | [66.7 - 67.5] |
| Canada | 67.1 | [66.7 - 67.5] | Lithuania | 66.6 | [66.2 - 67.0] |
| China | 69.3 | [68.9 - 69.7] | Luxembourg | 67.5 | [67.1 - 67.9] |
| Cyprus | 68.3 | [68.0 - 68.8] | Netherlands | 66.6 | [66.2 - 67.0] |
| Czech | 66.3 | [65.9 - 66.7] | Norway | 67.7 | [67.2 - 68.0] |
| Denmark | 66.5 | [66.1 - 66.9] | Portugal | 68.0 | [67.6 - 68.4] |
| Estonia | 67.2 | [66.7 - 67.5] | Romania | 68.0 | [67.6 - 68.3] |
| Finland | 64.8 | [64.3 - 65.2] | Slovakia | 67.0 | [66.6 - 67.4] |
| France | 66.2 | [65.8 - 66.6] | Slovenia | 65.3 | [64.9 - 65.7] |
| Germany | 65.8 | [65.4 - 66.2] | South Africa | 70.7 | [70.3 - 71.1] |
| Greece | 67.2 | [66.7 - 67.6] | Spain | 68.5 | [68.2 - 69.0] |
| Hungary | 66.9 | [66.5 - 67.4] | Sweden | 67.0 | [66.6 - 67.4] |
| India | 69.3 | [68.9 - 69.7] | Switzerland | 67.6 | [67.2 - 68.1] |
| Ireland | 69.2 | [68.9 - 69.7] | United States | 67.8 | [67.4 - 68.2] |
| Israel | 69.1 | [68.8 - 69.5] | United-Kingdom | 67.9 | [67.5 - 68.3] |

Table 4: DOIL value in each country with $R_0 = 2.5$.

| Country | DHIT (%) | | Country | DHIT (%) | |
|---|---|---|---|---|---|
| Australia | 41.2 | [38.3 - 44.4] | Italy | 40.5 | [37.7 - 43.5] |
| Austria | 41.3 | [38.5 - 44.1] | Japan | 42.4 | [39.4 - 45.0] |
| Bulgaria | 41.5 | [38.5 - 44.4] | Latvia | 42.5 | [39.5 - 45.2] |
| Canada | 42.2 | [39.2 - 45.1] | Lithuania | 42.0 | [39.0 - 44.9] |
| China | 42.5 | [39.6 - 45.3] | Luxembourg | 42.1 | [39.0 - 45.1] |
| Cyprus | 42.6 | [39.6 - 45.3] | Netherlands | 40.9 | [38.0 - 43.6] |
| Czech | 41.7 | [39.0 - 44.4] | Norway | 42.2 | [39.1 - 45.2] |
| Denmark | 42.0 | [39.2 - 45.0] | Portugal | 43.1 | [40.2 - 45.9] |
| Estonia | 42.1 | [39.0 - 44.8] | Romania | 41.0 | [38.1 - 43.9] |
| Finland | 41.3 | [38.1 - 43.9] | Slovakia | 42.2 | [39.1 - 45.0] |
| France | 41.1 | [38.1 - 44.0] | Slovenia | 42.4 | [39.2 - 45.2] |
| Germany | 40.9 | [37.7 - 43.6] | South Africa | 41.0 | [37.9 - 44.0] |
| Greece | 41.4 | [38.3 - 44.2] | Spain | 42.8 | [39.9 - 45.6] |
| Hungary | 40.3 | [37.6 - 43.2] | Sweden | 42.2 | [39.6 - 44.9] |
| India | 39.8 | [37.0 - 42.8] | Switzerland | 42.2 | [39.0 - 45.1] |
| Ireland | 43.4 | [40.6 - 46.1] | United States | 41.0 | [37.8 - 43.7] |
| Israel | 42.9 | [39.8 - 45.4] | United-Kingdom | 41.8 | [38.8 - 44.4] |

Table 5: DHIT value in each country with fixed transmissibility, so that $R_0 = 2.5$ in Italy.

| Country | DOIL (%) | | Country | DOIL (%) | |
|---|---|---|---|---|---|
| Australia | 67.4 | [67.0 - 67.8] | Italy | 66.3 | [65.9 - 66.7] |
| Austria | 67.1 | [66.7 - 67.5] | Japan | 68.6 | [68.2 - 69.0] |
| Bulgaria | 67.5 | [67.2 - 67.9] | Latvia | 68.7 | [68.3 - 69.1] |
| Canada | 68.8 | [68.3 - 69.2] | Lithuania | 68.3 | [67.9 - 68.6] |
| China | 69.3 | [68.9 - 69.7] | Luxembourg | 68.3 | [67.9 - 68.7] |
| Cyprus | 69.2 | [68.8 - 69.5] | Netherlands | 66.6 | [66.2 - 67.0] |
| Czech | 68.0 | [67.6 - 68.3] | Norway | 68.4 | [68.1 - 68.8] |
| Denmark | 68.1 | [67.8 - 68.6] | Portugal | 69.6 | [69.2 - 70.0] |
| Estonia | 68.0 | [67.6 - 68.3] | Romania | 67.2 | [66.7 - 67.6] |
| Finland | 67.3 | [66.9 - 67.7] | Slovakia | 68.6 | [68.2 - 69.0] |
| France | 67.0 | [66.6 - 67.4] | Slovenia | 68.6 | [68.2 - 69.0] |
| Germany | 66.6 | [66.2 - 67.0] | South Africa | 67.5 | [67.1 - 68.0] |
| Greece | 67.2 | [66.8 - 67.6] | Spain | 69.4 | [69.0 - 69.7] |
| Hungary | 66.1 | [65.7 - 66.6] | Sweden | 68.6 | [68.2 - 69.0] |
| India | 66.1 | [65.6 - 66.5] | Switzerland | 68.4 | [68.0 - 68.8] |
| Ireland | 70.1 | [69.6 - 70.4] | United States | 67.0 | [66.6 - 67.4] |
| Israel | 70.0 | [69.6 - 70.4] | United-Kingdom | 67.9 | [67.6 - 68.3] |

Table 6: DOIL value in each country with fixed transmissibility, so that $R_0 = 2.5$ in Italy.

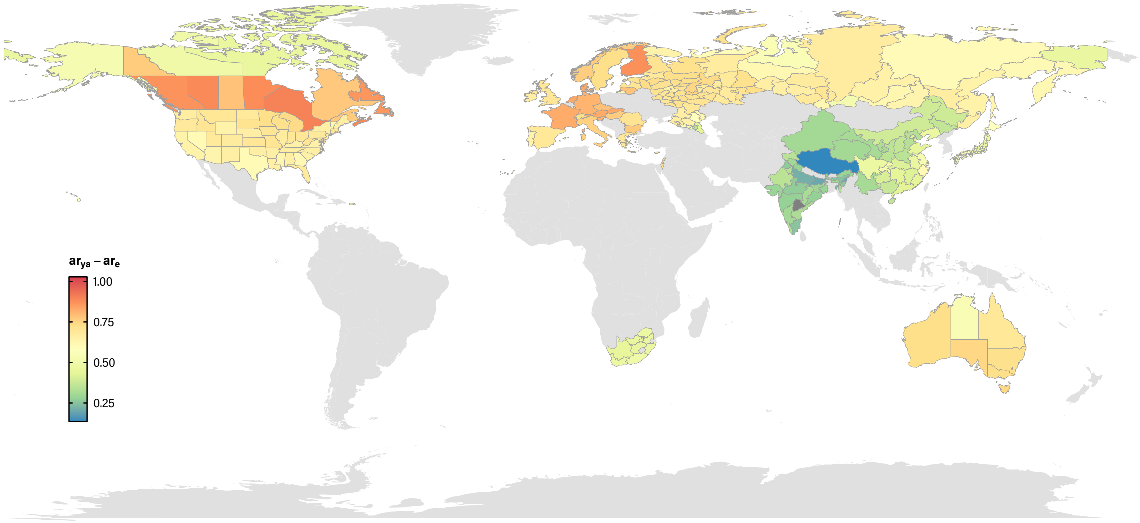

Figure 13: **Difference in the proportion of attack rates between young-adults and the elderly**. Regional variations between the attack rate proportion within the young-adults and the one for the elderly.

lower efficacy, the coverage must be larger than the DHIT value.

### 7 Further vaccination scenarios.

Besides the three vaccination strategies discussed in the main text, in this section we explore three other scenarios:

- Degree: vaccination is targeted towards nodes with the largest degree.
- Degree-uncorrelated: as in the previous strategy, but the network is rewired each day while preserving the degree of the nodes. Thus, age-age correlations are destroyed.
- Age Group: nodes are selected randomly from each age group with probability proportional to the average degree of the group. Nodes in the first 4 layers, representing individuals younger than 20 y.o. are not included.

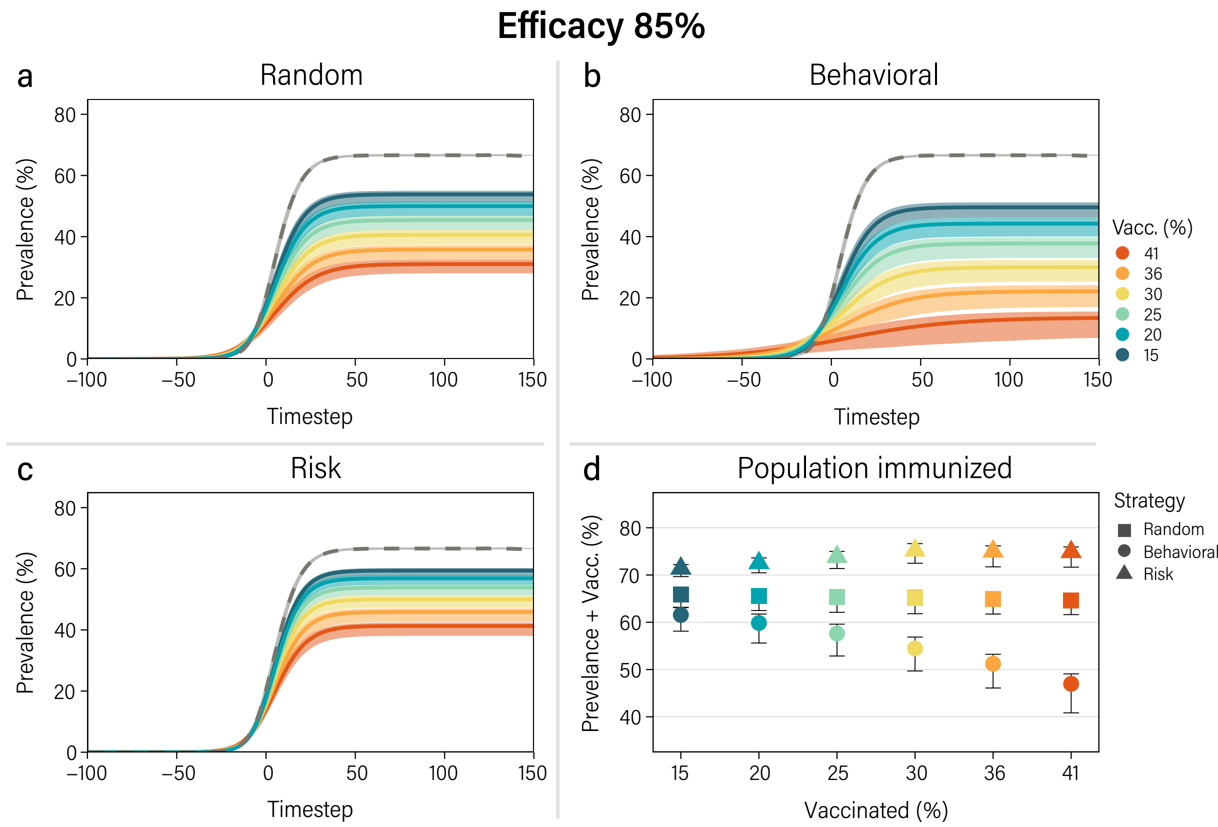

Figure 14: **Effect of different vaccination strategies.** Solid lines represent the median prevalence as a function of time under the random (a), behavioral-based (b) and risk-based (c) vaccine prioritization. The dashed lines account for a baseline scenario in which there is no vaccination nor any non pharmaceutical interventions. (d) Fraction of the population that is removed from the dynamics due to vaccine-induced immunity, disease-induced immunity or death once the disease dies out, for the different vaccination strategies as a function of the vaccine coverage. The results correspond to the population of Italy for $R_0 = 2.5$ and a vaccine efficacy of 85%.

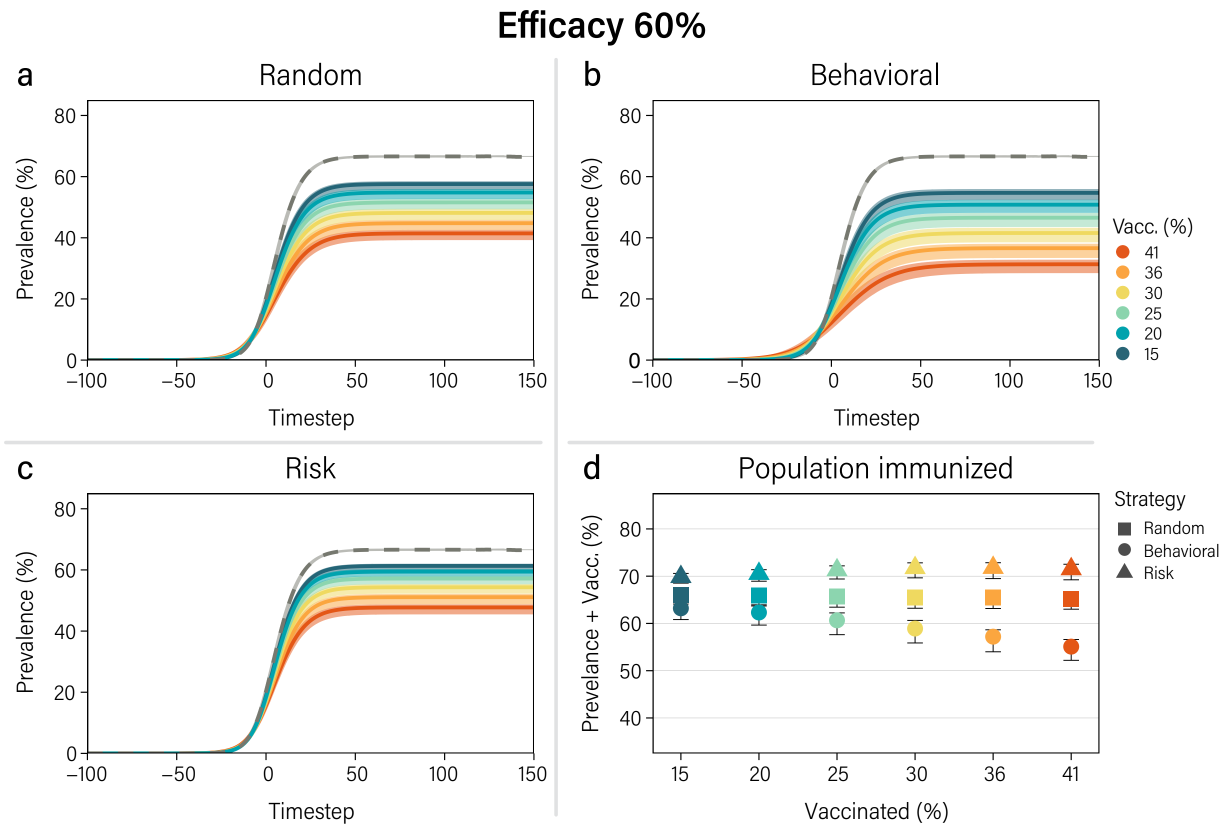

Figure 15: **Effect of different vaccination strategies.** Solid lines represent the median prevalence as a function of time under the random (a), behavioral-based (b) and risk-based (c) vaccine prioritization. The dashed lines account for a baseline scenario in which there is no vaccination nor any non pharmaceutical interventions. (d) Fraction of the population that is removed from the dynamics due to vaccine-induced immunity, disease-induced immunity or death once the disease dies out, for the different vaccination strategies as a function of the vaccine coverage. The results correspond to the population of Italy for $R_0 = 2.5$ and a vaccine efficacy of $60\%$.

| Number of averted deaths — $R_0$ = 2.5 — Vaccine efficacy = 85% | | | | | | |
|---|---|---|---|---|---|---|
| **Initially immune (%)** | **Random** | | **Behavioral** | | **Risk** | |
| 41 | 29.6 | [28.7 - 31.7] | 41.6 | [39.7 - 45.3] | 45.7 | [45.6 - 46.0] |
| 36 | 26.0 | [24.9 - 28.2] | 33.9 | [32.4 - 37.6] | 44.7 | [44.4 - 45.5] |
| 30 | 22.2 | [20.9 - 24.5] | 27.3 | [25.7 - 30.9] | 43.3 | [42.9 - 44.1] |
| 25 | 18.0 | [17.1 - 20.6] | 21.5 | [20.0 - 24.6] | 41.6 | [40.9 - 42.7] |
| 20 | 14.2 | [13.3 - 16.5] | 16.5 | [15.1 - 19.3] | 39.2 | [38.5 - 40.7] |
| 15 | 11.1 | [10.2 - 13.1] | 12.3 | [11.3 - 14.8] | 37.0 | [35.0 - 38.4] |

Table 7: Median number of averted deaths [95% C.I.] for each strategy with vaccine efficacy of 85%. In each strategy the population is initially immunized through vaccination under the rules explained in the main text.

| Number of averted deaths — $R_0$ = 2.5 — Vaccine efficacy = 60% | | | | | | |
|---|---|---|---|---|---|---|
| **Initially immune (%)** | **Random** | | **Behavioral** | | **Risk** | |
| 41 | 21.3 | [20.6 - 22.8] | 26.2 | [25.3 - 28.3] | 35.6 | [35.4 - 36.0] |
| 36 | 18.7 | [17.8 - 20.2] | 22.2 | [21.3 - 24.4] | 34.4 | [34.0 - 35.3] |
| 30 | 15.9 | [14.8 - 17.5] | 18.3 | [17.4 - 20.5] | 32.9 | [32.5 - 33.8] |
| 25 | 12.7 | [12.1 - 14.7] | 14.7 | [13.6 - 16.7] | 31.3 | [30.6 - 32.3] |
| 20 | 10.0 | [ 9.3 - 11.7] | 11.4 | [10.4 - 13.3] | 29.1 | [28.5 - 30.5] |
| 15 | 7.8 | [ 7.2 - 9.2] | 8.5 | [ 7.8 - 10.2] | 27.2 | [25.7 - 28.4] |

Table 8: Median number of averted deaths [95% C.I.] for each strategy with vaccine efficacy of 60%. In each strategy the population is initially immunized through vaccination under the rules explained in the main text.

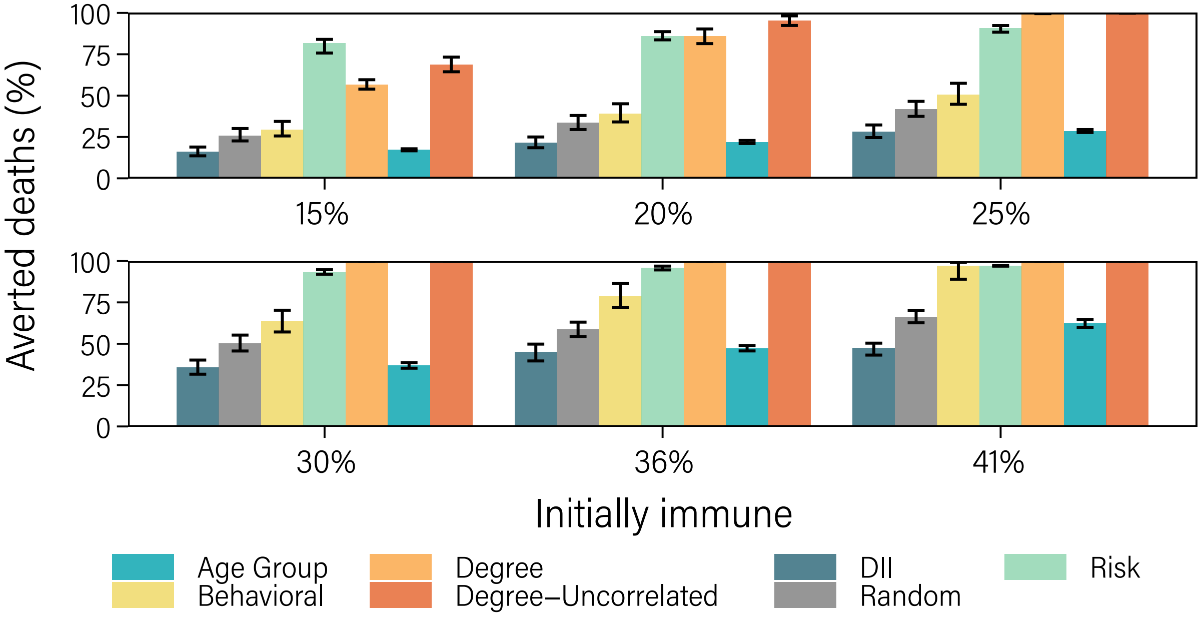

Figure 16: **Averted deaths in each vaccination scenario.** For each vaccination scenario we compute the percentage of deaths that would be averted in comparison to the baseline scenario ($y$-axis). The $x$-axis shows the initial fraction of immunized individuals at the beginning of the simulation.

In Fig. 16, we report the percentage of averted deaths in each scenario in comparison to the values computed for the baseline. We observe that for low vaccination coverage the risk based strategy results in the largest amount of averted deaths. On the other hand, for larger vaccination coverage the degree-based strategies outperforms the rest. However, identifying individuals with the largest number of connections is much harder in comparison to selecting individuals within the risk groups. Note also that the mechanism leading to the reduction of deaths is completely different in both scenarios. While in the risk-based strategy individuals are directly protected through vaccination, in degree-based ones they are protected thanks to the reduction of the overall incidence caused by the removal of highly connected nodes. This observation also explains why the age-group strategy is not so effective. Targeting groups with the largest degree excludes the elder groups from vaccination. Yet, since individuals are randomly chosen within the other groups, the coverage needed to reduce significantly the propagation of the disease is too large.
